# Transcriptional phenocopies of deleterious *KEAP1* mutations dictate survival outcomes in lung cancer treated with immunotherapy

**DOI:** 10.1101/2023.10.30.23297743

**Authors:** Stefano Scalera, Biagio Ricciuti, Daniele Marinelli, Marco Mazzotta, Laura Cipriani, Giulia Bon, Giulia Schiavoni, Irene Terrenato, Alessandro Di Federico, Joao V. Alessi, Maurizio Fanciulli, Ludovica Ciuffreda, Francesca De Nicola, Frauke Goeman, Giulio Caravagna, Daniele Santini, Ruggero De Maria, Federico Cappuzzo, Gennaro Ciliberto, Mariam Jamal-Hanjani, Mark M. Awad, Nicholas McGranahan, Marcello Maugeri-Saccà

## Abstract

Mutational models denoting KEAP1-NRF2 pathway activation have emerged as determinants of survival outcomes in non-small cell lung cancer (NSCLC). Hypothesizing that genetically distinct tumors recapitulate the transcriptional footprint of *KEAP1* mutations (KEAPness), we identified a KEAP1-NRF2-related gene set shared by tumors with and without pathway mutations. KEAPness-dominant tumors were associated with poor survival outcomes and immune exclusion in two independent cohorts of immunotherapy-treated NSCLC (SU2C and OAK/POPLAR). Moreover, patients with KEAPness tumors had survival outcomes comparable to their *KEAP1*-mutant counterparts. In the TRACERx421, KEAPness exhibited limited transcriptional intratumoral heterogeneity and an immune-excluded microenvironment, as highlighted by orthogonal methods for T cell estimation. This phenotypic state widely occurred across genetically divergent tumors, exhibiting shared and private cancer genes under positive selection when compared to *KEAP1*-mutant tumors. Collectively, we discovered the pervasive nature of the KEAPness phenotypic driver across evolutionary divergent tumors. This model outperforms mutation-based classifiers in predicting survival outcomes.

## Introduction

Immune checkpoint inhibitors targeting the programmed cell death protein 1/programmed death-ligand 1 (PD-1/PD-L1) pathway have reshaped the therapeutic landscape of advanced non-small-cell lung cancer (NSCLC) ^1^. Nevertheless, the limited accuracy of companion biomarkers in predicting clinical outcomes is a critical hurdle towards precision immuno-oncology. While immunohistochemical assessment of PD-L1 is routinely performed in the clinical setting, and generally correlates with increased benefit from PD-(L)1-based therapies, responses are also seen in tumors lacking PD-L1 expression ^1^. Similarly, tumor mutational burden (TMB) has been intensively investigated as a predictive biomarker. However, its use has not yet been implemented in clinical practice ^2^.

The Kelch-like ECH-associated protein 1 (KEAP1)-Nuclear factor erythroid-2-related factor 2 (NRF2) pathway is a core defensive mechanism against a variety of harmful cues, regulating cellular redox homeostasis and protecting cells against xenobiotics ^3–5^. In unstressed cells, KEAP1 triggers NRF2 proteasomal degradation through the CUL3-RBX1 E3 ubiquitin ligase complex. Oxidative and electrophilic stressors modify KEAP1 sensor cysteines, hampering its capability to bind the transcription factor NRF2. NRF2-driven transcription promotes metabolic rewiring and ensures protection against reactive oxygen species, xenobiotics, and ferroptosis ^3–5^.

*KEAP1* loss-of-function (LOF) mutations occur in approximately 15% of NSCLC ^6,7^, leading to uncontrolled NRF2 transcriptional activity and enhanced cytoprotection. In the specific context of non-squamous NSCLC, mostly consisting of lung adenocarcinoma (LUAD), mutations in the *KEAP1* tumor-suppressor gene are associated with resistance to chemotherapy, radiotherapy, and targeted agents, including KRAS-G12C inhibitors ^8–11^. To a similar extent, *KEAP1* mutations have been associated with inferior survival outcomes among non-squamous NSCLC patients treated with ICIs ^12–15^. Studies exploring the connection between *KEAP1* mutations and immunotherapy efficacy capitalized on the concept of immune-disrupting epistatic interactions, a genetic framework envisioning that cooperating genomic events elicit non-linear effects on cancer cell fitness and foster immune evasion. Consistently, co-existing *KEAP1* and *KRAS*/*STK11* mutations defined an uncommon subset of patients experiencing rapid disease progression during immunotherapy ^12,13^. Conversely, *KEAP1*/*TP53* co-mutations resulted in intermediate outcomes, denoting a neutral interaction ^14^. We have also described the deleterious nature of *KEAP1* mutations when associated with loss of heterozygosity (LOH) ^15^. Moreover, a connection between the magnitude of NRF2 transcription and *KEAP1*/*NFE2L2* mutation pathogenicity (damaging versus tolerated) was proposed ^16^. However, the same considerations do not extend to squamous cell lung carcinoma (LUSC). In this context, where *NFE2L2* (the gene encoding for NRF2) is more commonly mutated than *KEAP1* ^7^, evidence on the association between pathway mutations and inferior survival outcomes is still lacking. As a result, clinical outcomes of NSCLC patients receiving ICIs remain largely unpredictable beyond the limited fraction of patients whose tumors harbor specific mutational patterns, or that exhibit very high PD-L1/TMB levels.

We hypothesized that hallmarks of lethal *KEAP1* mutations are seeded in tumors with intact KEAP1-NRF2 pathway, and that this *KEAP1*-mutant-like state (KEAPness phenotype) shares the anti-immune properties of the canonical *KEAP1*-mutant counterpart, thus intercepting the large proportion of patients who do not obtain significant benefit from ICIs despite their tumors lack putative biomarkers of efficacy/resistance. On this basis, we conceived the following workflow: i) The TCGA was used for modelling KEAPness in NSCLC and to explore its association with the composition of the immune microenvironment at the pan-cancer level ^6,7,17,18^; ii) The impact of the KEAPness state on survival outcomes and immune phenotype was tested in two independent cohorts of metastatic NSCLC profiled by RNA-Seq and treated with ICIs: The Stand Up To Cancer-Mark Foundation (SU2C) NSCLC cohort (discovery cohort, n=153) and the OAK/POPLAR cohort (validation cohort, n=439) ^19–22^. This latter consists of patients enrolled in the phase II/III trials investigating second-line atezolizumab versus docetaxel in advanced NSCLC ^20,21^; iii) The prospective TRACERx421 multi-region sequencing study, which contains matched RNA-seq and whole-exome sequencing (WES) data from 347 early-stage NSCLC patients for a total of 947 tumor regions ^23^, was used to infer the evolutionary landscape of KEAPness tumors, the underlying oncogenic drivers, and KEAPness transcriptional heterogeneity. The workflow of the present study is illustrated in Figure 1.

**Figure 1.**
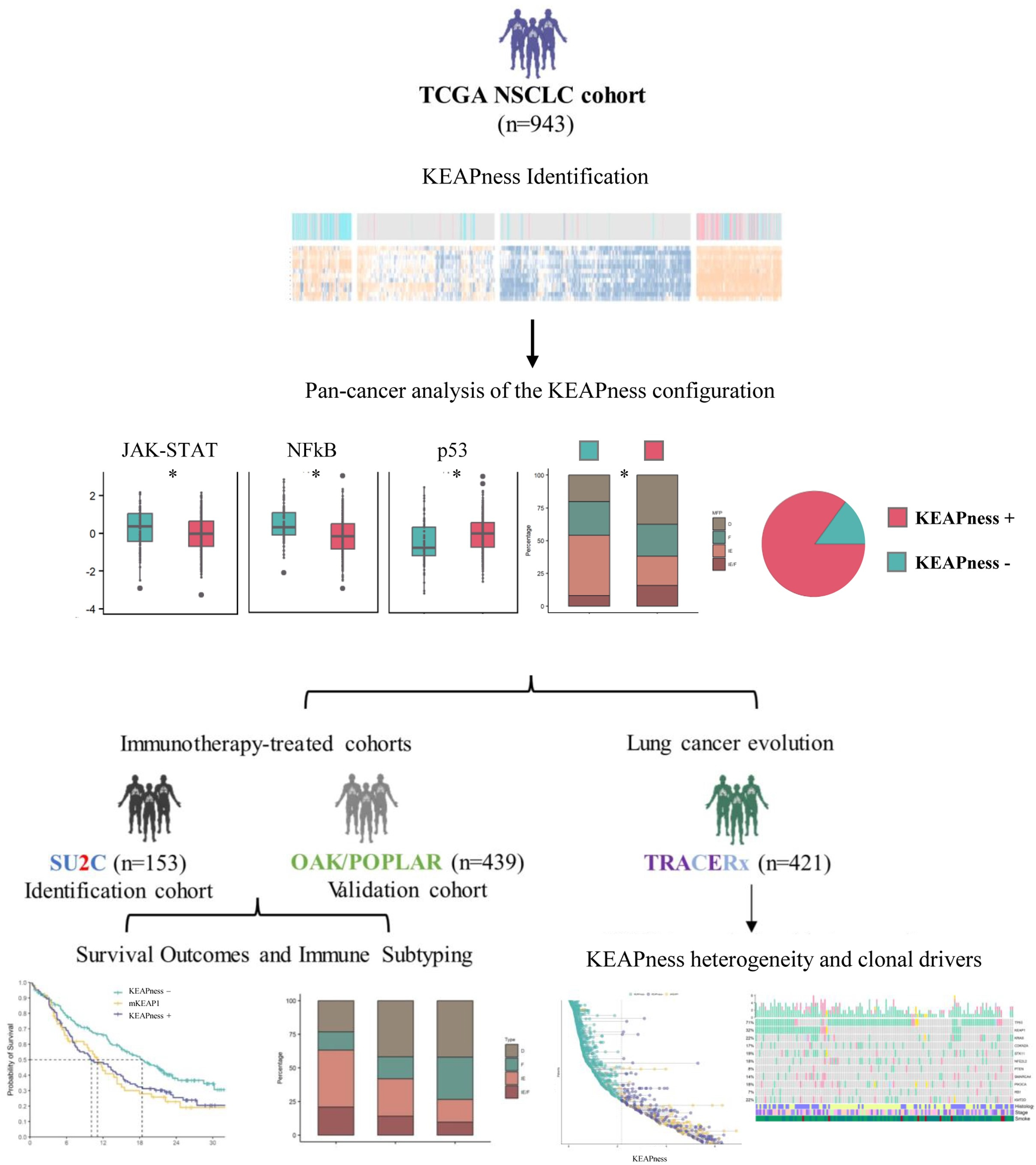
Study workflow. The TCGA NSCLC cohort was used to identify KEAP1-NRF2-associated genes shared by *KEAP1*-*NFE2L2*-mutant and wild-type cases. The relationship between the KEAPness configuration and the composition of the immune microenvironment was explored at the pan-cancer level (TCGA). The Stand Up To Cancer-Mark Foundation (SU2C identification cohort, n=153) and OAK/POPLAR (validation cohort, n=439) cohorts were used to investigate the impact of KEAPness on survival outcomes and immune subtyping in advanced NSCLC patients treated with immune checkpoint inhibitors. The TRACERx 421 multi-region sequencing study was exploited to investigate the distribution of KEAPness across evolutionary clusters, the underlying genetic drivers, and KEAPness transcriptional heterogeneity. This cohort was also used to validate the immunological features associated with KEAPness.

## Results

### Identification of the KEAPness state in the TCGA NSCLC cohort and pan-cancer analysis

To provide conceptual ground to our hypothesis that *KEAP1*-loss transcriptional phenocopies frequently occur in NSCLC, we first exploited an approach based on differential gene expression analysis between tumors with and without KEAP1-NRF2 pathway mutations, followed by unsupervised hierarchical clustering and gene co-expression. Evidence that a pool of positively correlated pathway core target genes are broadly expressed in NSCLC, regardless of the presence of *KEAP1* mutations, enabled us to distillate the KEAPness gene set (Figure 2a-c, Supplementary Table 1).

**Figure 2.**
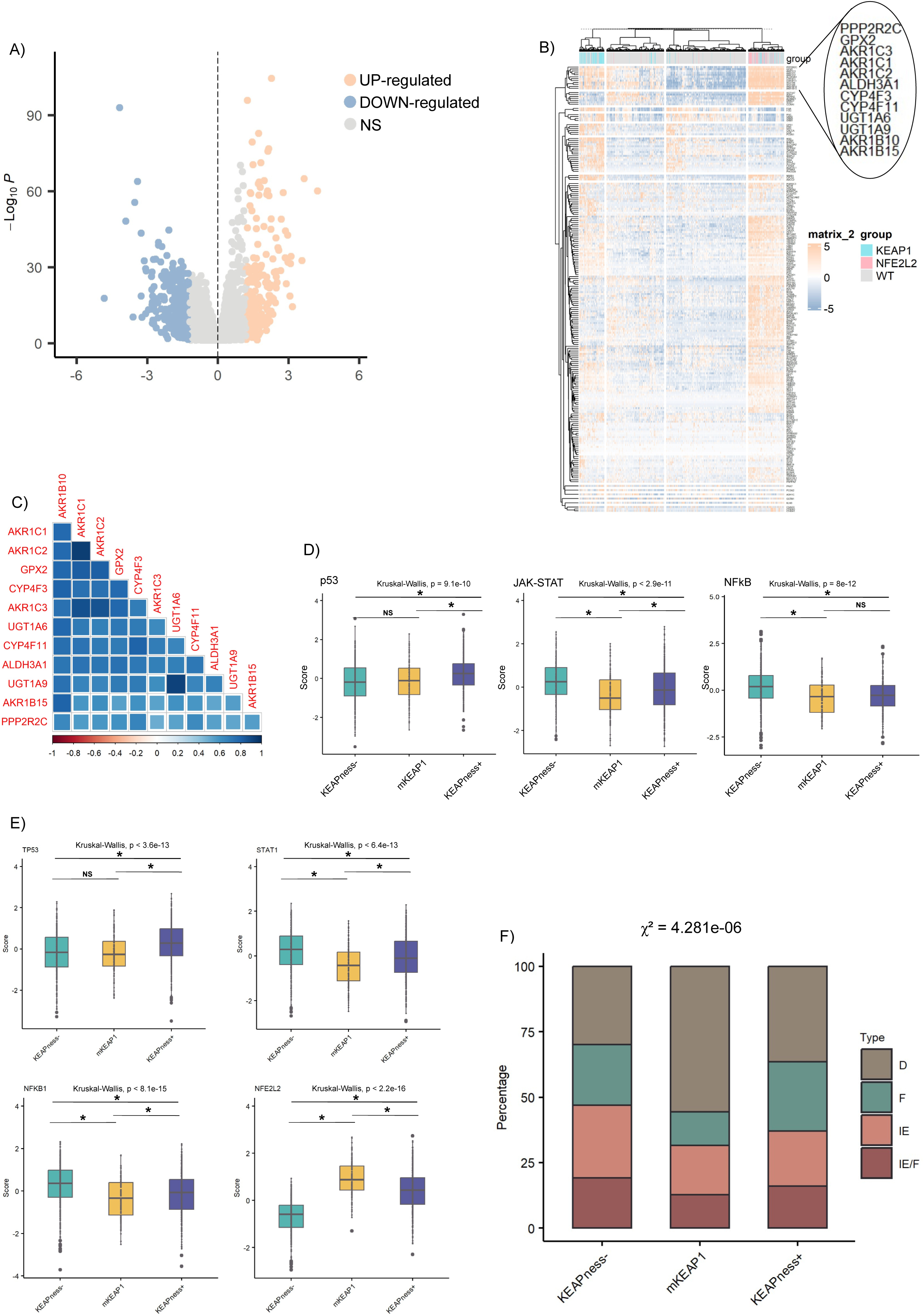
Identification of KEAPness in the TCGA NSCLC study. **a,** Volcano plot showing differentially expressed genes between *KEAP1*-*NFE2L2*-mutant and wild-type NSCLC (right: up-regulated, left: down-regulated; log2 fold change ≥1.2 and ≤ -1.2, adjusted p-value ≤ 0.05). **b,** Heatmap showing unsupervised hierarchical clustering of differentially expressed genes, with magnification of the gene set of interest. **c,** Heatmap illustrating Pearson correlation coefficients for the KEAPness genes. **d,** Box plots illustrating the differences in the p53, JAK-STAT, and NFkB PROGENy signatures across KEAPness (KEAPness+), *KEAP1*-mutant (mKEAP1) and KEAPness-free (KEAPness-) tumors. **e**, Box plots illustrating DoRothEA-predicted activity of selected transcription factors in the same subsets. **f,** Stacked bar chart displaying the distribution of immune subtypes. D: desert, immune-depleted and non-fibrotic, F: fibrotic, immune-depleted and fibrotic, IE: immune-enriched and non-fibrotic, IE/F: immune-enriched and fibrotic.

To better characterize this transcriptional configuration, we investigated its relationship with histology, cancer-associated pathways inferred from perturbation experiments (PROGENy) ^24^, and activity of immune-and pathway-related transcription factors (DoRothEA) ^25^. We noticed that LUSC had a significantly higher frequency of KEAPness when compared to LUAD (Supplementary Figure 1). Moreover, KEAPness tumors exhibited a distinct profile of pathway-level gene expression signatures (e.g., p53 and JAK-STAT) compared to KEAPness-free samples (Figure 2d; and Extended Data Figure 1). Differences in the related transcription factors were confirmed by estimating transcription factor-target interactions (Figure 2e). Next, we exploited immune subtyping from bulk RNA-Seq to more accurately examine the tumor immune microenvironment associated with the KEAPness state ^26^. We noted that NSCLC with KEAPness had an immune-excluded microenvironment, which was comparable to that of tumors with *KEAP1* mutations (Figure 2f; and Extended Data Figure 2).

To add a further level of resolution to these observations, we explored the distribution of the *KEAP1*-mutant-like phenotype, and its immunological correlates, across ∼ 8,000 *KEAP1* wild-type tumors samples spanning 17 cancer types (Figure 3, and Supplementary Figure 2). Consistently with results from NSCLC, we observed that KEAPness was enriched in squamous tumors, such as head and neck squamous cell carcinoma, esophageal cancer and cervical cancer. In these tumors, KEAPness exhibited an immunological background and repertoire of pathway-level gene expression signatures which are reminiscent of NSCLC. To a similar extent, bladder cancer, colorectal cancer and gastric adenocarcinoma frequently carried the KEAPness configuration, which was accompanied by similar immunological and gene expression features. Next, KEAPness was extremely common in hepatocellular carcinoma, a finding that is consistent with the detoxification mechanisms mediated by the KEAP1-NRF2 pathway. Conversely, hormone-dependent tumors (e.g., breast and prostate cancers), gynecologic tumors (e.g., endometrial and ovarian cancers), and non-epithelial cancers (glioblastoma, skin melanoma, and sarcomas) were prevalently KEAPness-free.

**Figure 3.**
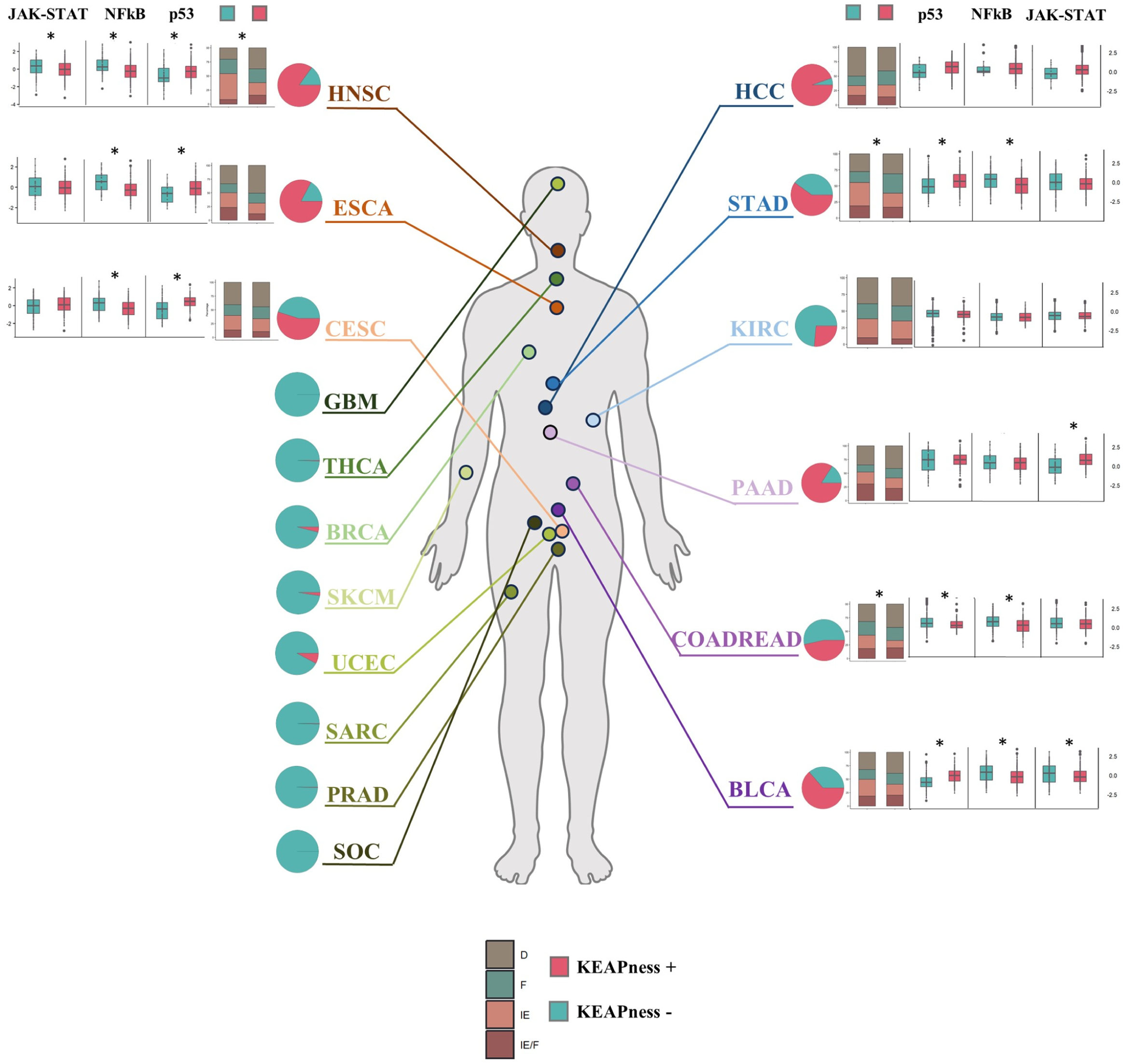
Frequency of KEAPness at the pan-cancer level (TCGA). Distribution of KEAPness across 17 *KEAP1* wild-type cancer types, along with the associated immune subtyping and selected pathway signatures. Abbreviations: HNSC: head and neck squamous cell carcinoma, ESCA: esophageal carcinoma, CESC: cervical squamous cell carcinoma, GBM: Glioblastoma, THCA: thyroid carcinoma, BRCA: breast cancer, SKCM: skin cutaneous melanoma, UCEC: uterine corpus endometrial carcinoma, SARC: sarcoma, PRAD: prostate adenocarcinoma, SOC: serous ovarian carcinoma, HCC: hepatocellular carcinoma, STAD: stomach adenocarcinoma, KIRK: kidney renal clear cell carcinoma, PAAD: pancreatic adenocarcinoma, COADRED: colorectal cancer, BLCA: bladder cancer.

Overall, the immunological background associated with KEAPness, denoting immune exclusion, closely resembles that of *KEAP1*-mutant NSCLC. Moreover, pan-cancer analyses revealed that this phenotypic state is a core feature of squamous tumors which also sporadically affects other histological types.

### KEAPness enrichment predicts survival outcomes and immunotherapy efficacy in two independent clinical cohorts

Having defined that KEAPness is a common phenomenon in NSCLC, we next assessed whether a parsimonious binary model exclusively built on the KEAPness gene set predicted clinical outcomes among advanced NSCLC patients treated with immunotherapy. Using the receiver operator characteristic (ROC) curve and bootstrap resampling for predicting a lack of clinical benefit (disease progression within six months from the beginning of ICIs), we defined the most accurate KEAPness cut point in the SU2C identification cohort (Extended Data Figure 3). To avoid confounding factors related to histology and previous therapies, this analysis was carried out in non-squamous NSCLC patients who did not receive prior tyrosine kinase inhibitors (TKIs) (n=93). This threshold was used for all subsequent survival analyses and immune subtyping in the two independent clinical cohorts (SU2C and OAK/POPLAR), as well as the TRACERx 421 cohort. Baseline clinical characteristics of patients included in the SU2C and OAK/POPLAR cohorts, and their association with the phenotype investigated, are reported in Supplementary Table 2-4.

In the SU2C cohort (n=153), patients with KEAPness-dominant tumors (i.e., those with a gene expression profile compatible with a KEAPness state when *KEAP1* mutations are not considered in the model) had shorter progression-free survival (PFS) and overall survival (OS) compared to those with KEAPness-free tumors (PFS log-rank p=0.042; OS log-rank p=0.008) (Figure 4a-b; Extended data figure 4a-b). Likewise, progressive disease (PD)/stable disease (SD) and immune exclusion were more frequently observed among KEAPness-dominant tumors (Figure 4c-d, and Supplementary Figure 3a). PD-L1 expression levels were lower among KEAPness-dominant NSCLC, even though this difference was not significant due to the limited size of the subgroups compared (n=71; Supplementary Figure 4). Considering the limited role of ICIs in oncogene-addicted NSCLC, we carried out a sensitivity analysis upon exclusion of patients who received prior TKIs. In this case, the performances of the classifier were further improved (Supplementary Figure 5).

**Figure 4.**
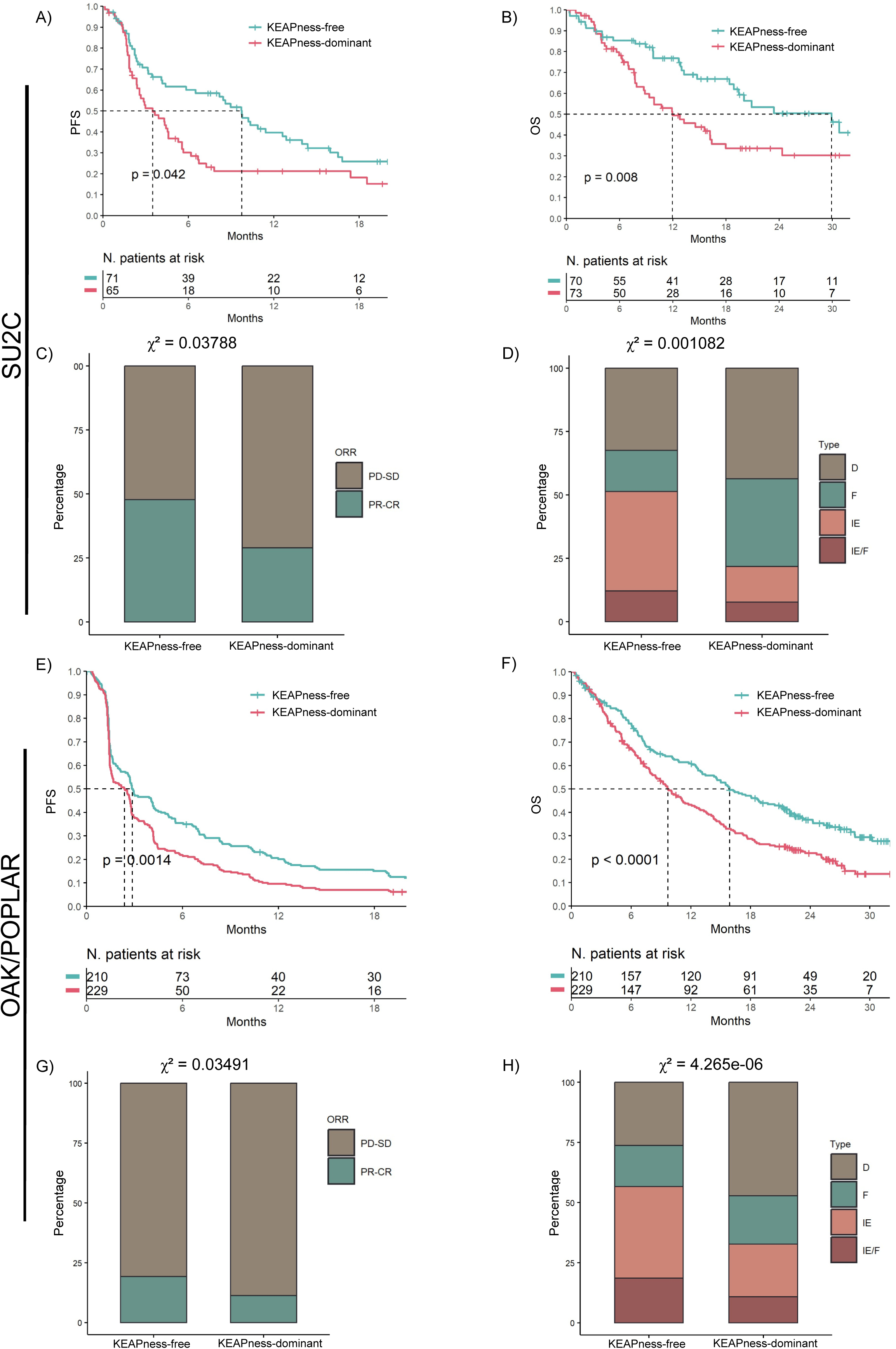
Survival analyses and immune subtyping in the SU2C identification cohort and OAK/POPLAR validation cohort. **a, b**, Kaplan-Meier survival curves for progression-free survival (PFS) and overall survival (OS) comparing KEAPness-dominant and KEAPness-free NSCLC in the SU2C cohort. **c,** Stacked bar chart illustrating the differences in the overall response rate (ORR) between patients with KEAPness-dominant and KEAPness-free NSCLC in the SU2C study. Abbreviations: CR: complete response, PR: partial response, SD: stable disease, PD: progressive disease. **d,** Stacked bar chart showing the distribution of immune subtypes in the SU2C cohort. Abbreviations: D: desert, immune-depleted and non-fibrotic, F: fibrotic, immune-depleted and fibrotic, IE: immune-enriched and non-fibrotic, IE/F: immune-enriched and fibrotic. **e-h,** Survival analyses (e, f), ORR (g), and immune subtypes (h) in the OAK/POPLAR validation cohort.

Results were entirely confirmed in the independent OAK/POPLAR validation cohort (atezolizumab, n=439) in terms of survival outcomes, immunological correlates, and frequency of the investigated phenotype (PFS log-rank p=0.0014; OS log-rank p<0.0001) (Figure 4e-h; Extended data figure 4c-d, and Supplementary Figure 3b). Of note, KEAPness-dominant tumors were associated with impaired clinical outcomes also in a separate cohort of chemotherapy-treated patients in the OAK/POPLAR trials (docetaxel, n=452; Supplementary Figure 6).

Further corroborating the consistency of our results, we confirmed that KEAPness-dominant tumors were enriched for the squamous histology (Supplementary Figure 7). However, the clinical implications of the KEAPness-dominant phenotype were independent of histology, even though survival analyses in squamous tumors were limited by the relatively low sample size (Supplementary Figure 8).

Collectively, these data indicate that a streamlined RNA-only classifier frames a large population of patients with tumors exhibiting *KEAP1*-mutant-like traits, and efficiently predicts immunotherapy efficacy and immune exclusion in advanced NSCLC patients. This classifier divides the population in two groups of comparable size, thus indicating that the model does not exclusively identify outlier patients.

### Patients with KEAPness and *KEAP1*-mutant tumors have similar survival outcomes when treated with immunotherapy

Having validated a model exclusively based on gene expression in the two independent cohorts, we used the pooled SU2C/OAK/POPLAR cohort with matched RNA-Seq and WES data to unambiguously demonstrate that KEAPness tumors recapitulate the canonical *KEAP1*-mutant counterparts. To this end, we compared the predictive ability of a combined model that included *KEAP1* mutations in the gene expression classifier to exactly frame the “pure” KEAPness configuration. Data on *KEAP1* mutations were available from 61 patients in the SU2C cohort and 251 patients in the OAK/POPLAR cohort.

In survival analyses, we confirmed the hypothesis that patients with KEAPness had shorter PFS and OS compared to those with KEAPness-free tumors (Figure 5a-b). Importantly, clinical outcomes of the KEAPness group were comparable to those of the *KEAP1*-mutant group (Figure 5a-b). A comparable pattern was observed when considering tumor radiological response to immunotherapy (Figure 5c). KEAPness was significantly more common that *KEAP1* mutations, being observed in approximately 30% of the whole population, thus formally confirming the hypothesis that a model based on gene expression outperforms mutational classifiers in predicting immunotherapy efficacy. Next, we validated in the clinical cohorts that NSCLC with KEAPness more frequently exhibited an immune-excluded microenvironment compared to KEAPness-free tumors, having an immunological phenotype reminiscent of that of *KEAP1*-mutant tumors (Figure 5d-e). The same considerations extend to p53 and JAK-STAT gene expression signatures (Extended Data Figure 5).

**Figure 5.**
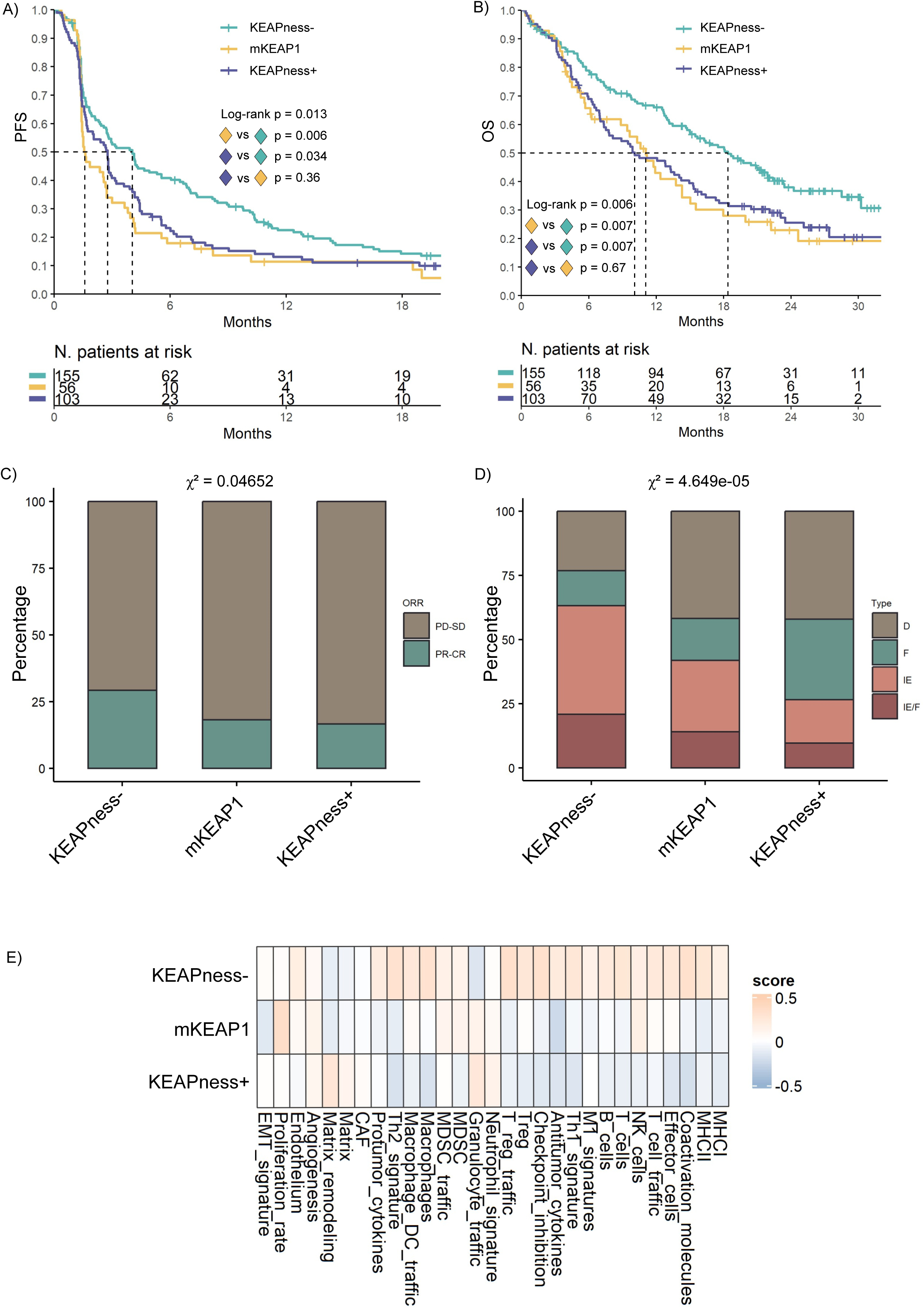
Survival analyses in the pooled SU2C/OAK/POPLAR cohort to explore the clinical significance of KEAPness in relation to *KEAP1* mutations. **a, b,** Kaplan-Meier survival curves of progression-free survival (PFS) and overall survival (OS) comparing KEAPness (KEAPness+), *KEAP1*-mutant (mKEAP1), and KEAPness-free (KEAPness-) NSCLC. **c,** Stacked bar chart displaying the differences in terms of overall response rate (ORR) across the subgroups compared. **d,** Stacked bar chart for immune subtyping across the three molecular subgroups considered. **e,** Heatmap illustrating the 29 tumor immune microenvironment (TIME) signatures used for immune subtyping (BostonGene) across the three subgroups compared. Abbreviations: D: desert, immune-depleted and non-fibrotic, F: fibrotic, immune-depleted and fibrotic, IE: immune-enriched and non-fibrotic, IE/F: immune-enriched and fibrotic.

Overall, while a binary RNA-based model identified a large proportion of NSCLC that are poorly responsive to ICIs, we confirmed that KEAPness and *KEAP1*-mutant tumors share comparable survival outcomes and immune-related features.

### Genetic selection and evolutionary trajectories in the TRACERx 421 cohort

We next investigated the genetic and evolutionary correlates of KEAPness leveraging multi-region WES and transcriptome sequencing data from the TRACERx 421 cohort (previously untreated, early-stage NSCLC) ^23, 27^.

First, we investigated genetic selection among more than 700 cancer genes using the dNdScv method^28^. We identified a set of driver genes under positive selection (q-value < 0.05), with differences in relation to histology (Figure 6a-b). In non-squamous NSCLC, KEAPness and *KEAP1*-mutant tumors shared positive selection for *TP53*, *KRAS* and *STK11* (Figure 6a). However, while *SMARCA4* was under positive selection exclusively in the *KEAP1*-mutant group, *CDKN2A* (p14(ARF)) was identified as a private driver in non-squamous NSCLC with KEAPness (Figure 6a). In squamous tumors, *TP53* and *CDKN2A* were identified as shared drivers between *KEAP1*-mutant and KEAPness tumors (Figure 6b). Conversely, squamous tumors with KEAPness exhibited positive selection for PI3K pathway genes (*PIK3CA* and *PTEN*) and *NFE2L2* (Figure 6b). Most non-silent mutations in these cancer-associated genes were of clonal nature (Figure 6c). We also observed a similar number of clonal non-synonymous mutations and a similar frequency of genome doubling across *KEAP1*-mutant, KEAPness, and KEAPness-free tumors (Figure 6d-e), indicating that clinical findings are unrelated to these events.

**Figure 6.**
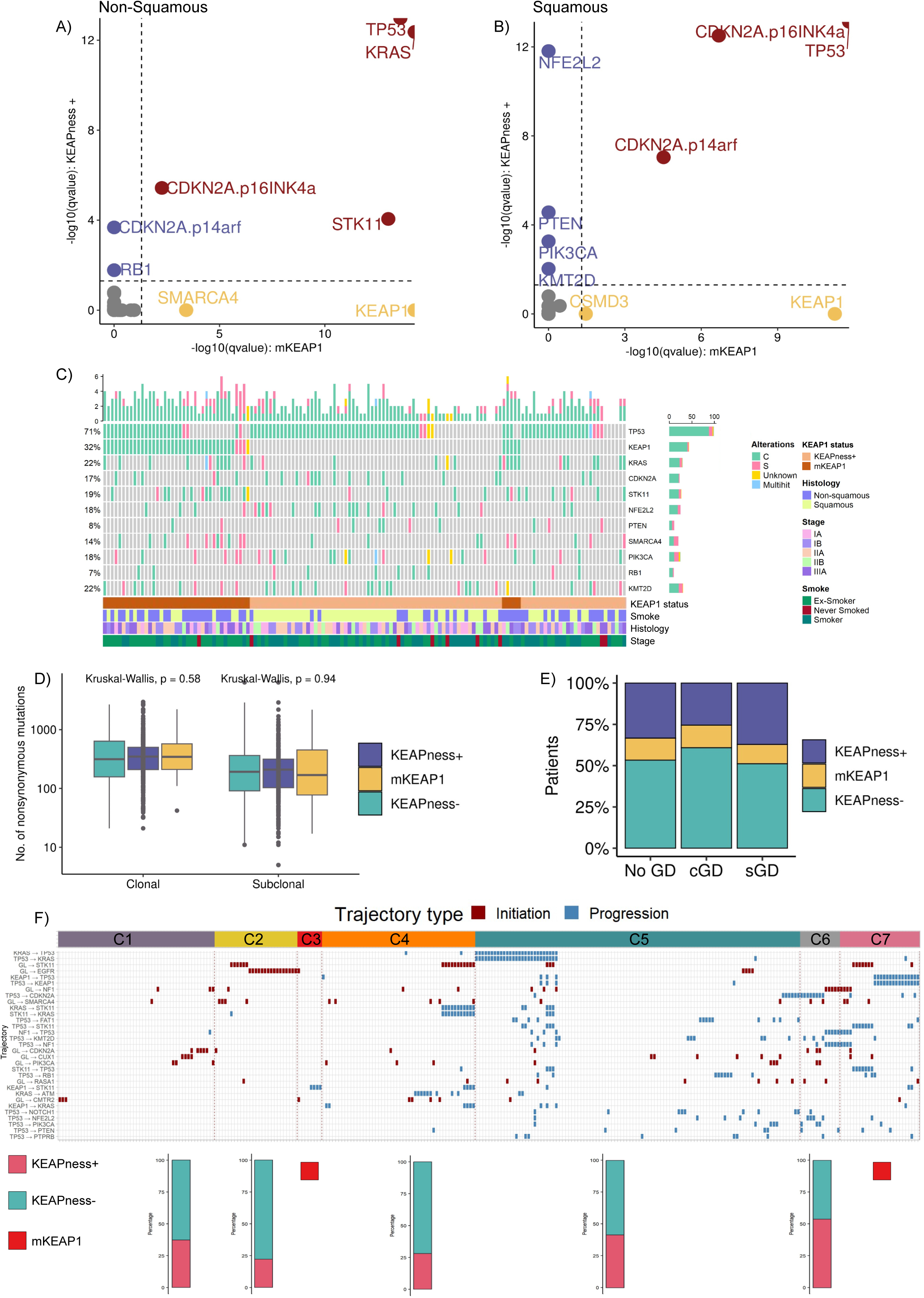
Genetic selection and evolutionary trajectories in the TRACERx 421 cohort. **a, b,** Shared and private COSMIC cancer genes under positive selection in KEAPness and *KEAP1*-mutant non-squamous (a) and squamous (b) tumors (red: shared genes; blue: private genes in KEAPness tumors; yellow: private genes in *KEAP1*-mutant tumors). **c,** Oncoprint illustrating the mutational frequency and clonal status of genes under positive selection in KEAPness and *KEAP1*-mutant NSCLC. Clonal mutations are indicated in water green. **d**, **e,** Number of clonal and subclonal non-synonymous mutations in KEAPness (KEAPness+), *KEAP1*-mutant (mKEAP1) and KEAPness-free (KEAP1ness-) tumors (d) and presence of clonal and subclonal genome doubling (cGD and sGD). **f**, Oncoprint for repeated evolution (REVOLVER) showing the distribution of the KEAPness phenotype across genetically divergent clusters (bottom), and the driver trajectories detected in each cluster (left side of the oncoprint). The trajectory type is also reported, e.g., initiation: germline (GL)➔*TP53*; progression: *KRAS*➔*STK11*.

We then assessed the relationship between evolutionary trajectories of driver alterations and the KEAPness phenotype using REVOLVER, a method that exploits transfer learning to infer repeated evolution by highlighting hidden evolutionary patterns (Figure 6f) ^29^. We discovered that KEAPness pervasively occurred across multiple evolutionary trajectories. The lowest frequency was noted in *EGFR*-driven tumors, a finding that is consistent with the different etiology of *KEAP1*-and *EGFR*-mutant NSCLC (smoking exposure).

Thus, KEAPness exhibits both private and shared cancer genes under positive selection when compared to the *KEAP1*-mutant disease, and intercepts genetically divergent tumors with a similar clinical course and response to immunotherapy.

### KEAPness transcriptional heterogeneity and immunological correlates in the TRACERx 421 cohort

We further exploited the TRACERx 421 to investigate the heterogeneity of the KEAPness state and expand the study of the associated immunological features.

First, we observed that KEAPness was a stable phenotype across multiple regions, given that only 31% of KEAPness tumors had at least one region classified as KEAPness-free (Figure 7a). This limited intratumoral heterogeneity denotes a phenotype which is consistent across time and space in tumor evolution. However, non-squamous NSCLC had a higher fraction of KEAPness heterogeneous tumors when compared to squamous NSCLC, thus further highlighting the predominant role of KEAPness in the squamous histology (Extended Data Figure 6).

**Figure 7.**
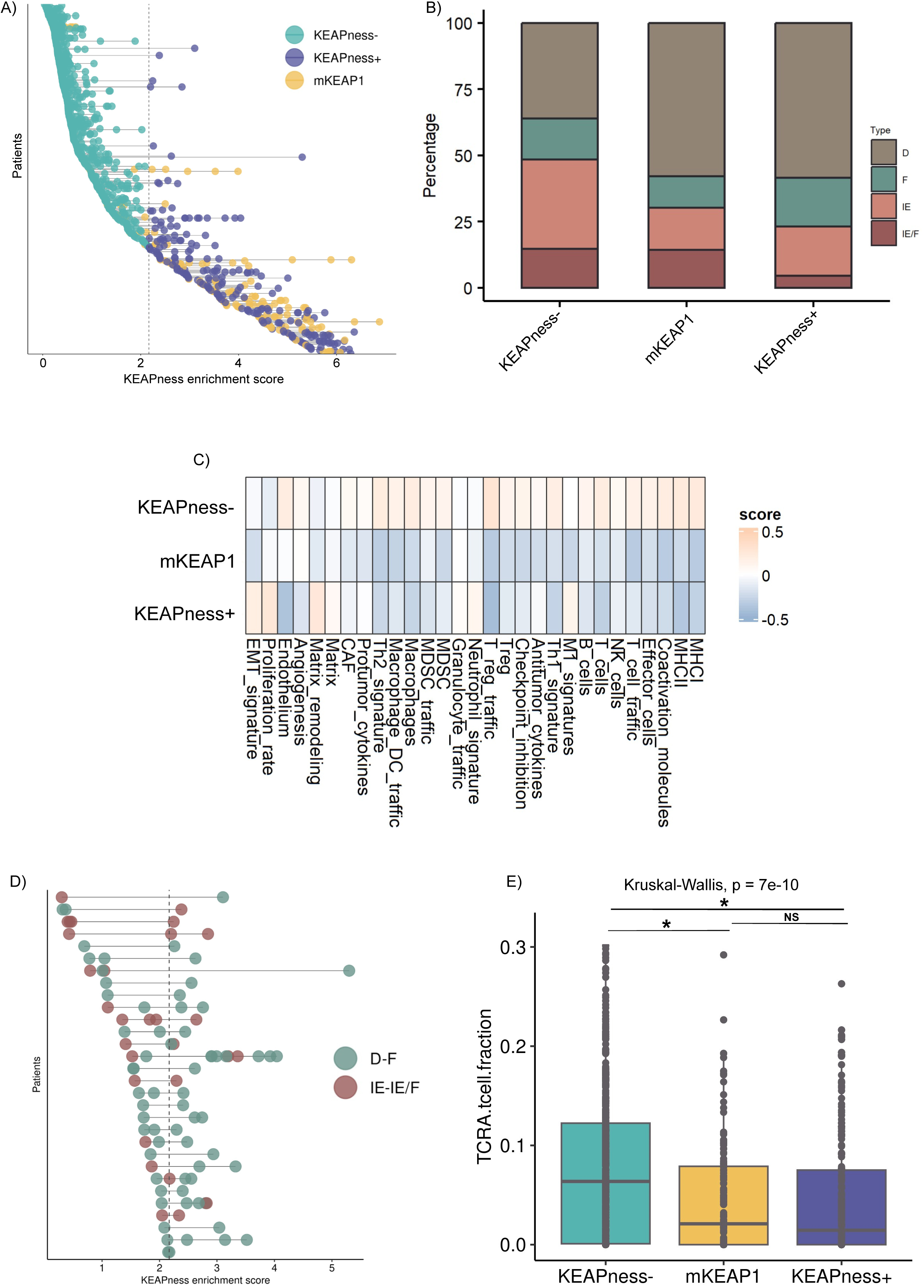
KEAPness intratumoral heterogeneity and immunological correlates in the TRACERx 421 cohort. **a,** Dumbbell plot showing KEAPness at the patient-level. Each dot represents a tumor region. The dotted line represents the KEAPness cutoff identified in clinical cohorts. Tumors with KEAPness transcriptional heterogeneity are those with both water green and blue dots. **b,** Stacked bar chart displaying the sample-level distribution of the four immune subtypes across KEAPness (KEAPness+), *KEAP1*-mutant (mKEAP1), and KEAPness-free (KEAPness-) NSCLC. D: desert, immune-depleted and non-fibrotic, F: fibrotic, immune-depleted and fibrotic, IE: immune-enriched and non-fibrotic, IE/F: immune-enriched and fibrotic. **c,** Heatmap illustrating the 29 TIME signatures exploited for generating four immune subtypes (BostonGene) across the same subgroups. **d,** Dumbbell plot showing KEAPness heterogeneous tumors and the related region-level immune subtyping (D-F: desert/fibrotic, IE-IE/F immune-enriched, either fibrotic or not). **e,** Box plot illustrating T cell infiltration estimated by T cell ExTRECT.

We then explored the immunological features associated with KEAPness. As already reported in the TCGA and in clinical cohorts, NSCLC with KEAPness were more commonly immune-excluded compared to KEAPness-free tumors, resembling the *KEAP1*-mutant counterparts (Figure 7b-c). Among KEAPness heterogeneous tumors, namely, KEAPness tumors with at least one KEAPness-free region, we observed a mixed immune-related pattern (Figure 7d). While some tumors were immunologically stable having the same immune subtype in all the tumor regions, in other tumors the acquisition of KEAPness was paralleled by changes in the tumor microenvironment (i.e., from immune-enriched to immune-excluded), suggesting tumor-microenvironment co-evolution (Figure 7d). Lastly, we leveraged T cell ExTRECT, a method that infers T cell infiltration from DNA sequencing, rather than RNA-Seq, for orthogonal validation of immune-related features ^30^. KEAPness tumors had lower T-cell infiltration compared to the KEAPness-free counterparts, being comparable to *KEAP1*-mutant tumors (Figure 7e).

Collectively, these analyses conveyed the message that KEAPness is characterized by limited intratumoral heterogeneity, and provided unbiased evidence on the relationship between this novel cancer phenotype and the immune microenvironment considering the reproducibility of the data across multiple independent cohorts and with different methods.

## Discussion

Accumulating evidence links *KEAP1*-based mutational contexts to survival outcomes in NSCLC patients treated with immunotherapy ^12–15^. However, dissecting the impact of *KEAP1* mutations on immunotherapy outcomes is challenging, given that co-mutations in *KEAP1*/*KRAS*/*STK11* occur in approximately 10-15 % of non-squamous NSCLC patients ^12, 13^. In addition, these models cannot be extended to squamous tumors, which have a distinct genomic profile characterized by *TP53* alterations and lack of *KRAS*/*STK11* mutations ^7^. Biomarkers associated with DCB represent the other side of the challenging task of predicting ICIs efficacy, considering the limited accuracy of PD-L1 and TMB. High PD-L1 levels (≥ 50%) are detected in approximately 30% of NSCLC ^31^, and are associated with greater benefit from immunotherapy. However, primary and acquired resistance are common among patients with PD-L1-expressing NSCLC. Similarly, tumor responses are often observed in PD-L1 low/negative NSCLC. The use of TMB has also been difficult to implement in clinical practice. Beyond the discrepancies among the various pipelines ^2^, we have witnessed a wealth of studies which progressively raised the TMB cut-off, from ≥ 10 non-synonymous mutations per megabase (mut/Mb) to more extreme values ^2, 32–36^. Intuitively, while raising the TMB is expected to improve its predictive ability, very high TMB thresholds significantly lower the TMB-high population, which drops down to 10-15% of the whole population ^36^. Thus, there is a pressing need for improved biomarkers which can predict ICIs efficacy.

*KEAP1* LOF mutations lead to aberrant NRF2 transcriptional activity. Here, we hypothesized that analyzing the KEAP1/NRF2 pathway from a transcriptional perspective may help overcome the many limitations of current mutation-based biomarkers by identifying transcriptional phenocopies of driver *KEAP1* mutations. By integrating survival analyses and computational studies, we reported that: i) Transcriptional phenocopies of *KEAP1* mutations are common in NSCLC and across a variety of cancer types, particularly squamous tumors; ii) A gene expression model denoting the transcriptional output of *KEAP1* mutations predicts inferior survival outcomes, lower response to immunotherapy, and immune exclusion, regardless of the presence of *KEAP1* mutations; iii) KEAPness phenodrivers have limited, but not negligible, transcriptional heterogeneity and associate with immune exclusion. Furthermore, KEAPness occurs across genetically divergent tumors and, when compared to the *KEAP1*-mutant disease, exhibits some private genomic traits driving tumor evolution. Our study has important strengths. First, KEAPness was identified in the TCGA NSCLC cohort, and then applied to the clinical cohorts. Second, we used the same classification for survival analyses, which yielded comparable results in terms of survival outcomes, distribution of cases, and immunological correlates in independent cohorts. Likewise, the same cut-off was also applied to the TRACERx 421, where immunological features of the KEAPness configuration were comparable to those observed in the other cohorts, and further confirmed by a different method leveraging DNA sequencing (T cell ExTRECT) ^30^. Third, we used a streamlined method to define KEAPness-dominant tumors, which could be easily exploited in the clinical setting. Indeed, while we dissected the clinical, biological and evolutionary patterns of KEAPness tumors, a parsimonious binary model for outcome prediction is largely preferred from a clinical perspective. Importantly, we pursued an identification-validation approach, evaluating data from patients enrolled into a randomized phase III trial (OAK). In the search for cancer biomarkers, these represent important methodological added values. The distribution of KEAPness-dominant cases deserves further mention. While current biomarkers mostly focus on outlier patients, the split achieved by our model suggests that it better delineates the actual population of immunotherapy responsive/resistant NSCLC.

From a clinical perspective, this study has several implications. Multiple first-line treatment options are available for patients with metastatic non-small cell lung cancer, including PD-(L)1 monotherapy alone or in combination with CTLA-4 inhibitors and/or chemotherapy. Nevertheless, the lack of robust biomarkers is a critical hurdle in clinical practice. Our data suggest that patients with KEAPness tumors have worse outcomes to PD-(L)1-based monotherapies, and that intensifying treatment with the addition of CTLA-4 blockade or chemotherapy may be a reasonable strategy, considering that the addition of CTLA-4 inhibitors and chemotherapy can be beneficial in immune-excluded tumors with low/negative PD-L1 expression ^37–39^. Importantly, an increasing number of commercial assays are now exploiting WES and RNA-seq, highlighting how gene expression signature can be incorporated in clinical decision making. Our data also have implications for patients with early-stage NSCLC. PD-(L)1 blockade is now approved as adjuvant and neoadjuvant therapy ^40–42^. However, the only marker of improved long-term outcomes is pathologic complete response in the case of pre-operative immunotherapy. Indeed, PD-L1 expression and TMB have been inconsistently associated with the risk of recurrence in these settings. Therefore, we envision opportunities for the use of additional biomarkers to identify early-stage NSCLC patients who will benefit from intensification of perioperative treatment.

Although our data were consistent between two independent clinical cohorts, we acknowledge that our study has some limitations. The main limitation is a lack of data on PD-L1 and TMB, apart from a subset of patients in the SU2C cohort. Thus, we were unable to include these features in multivariate Cox regression models. Likewise, a limited number of clinical parameters were available in the OAK/POPLAR cohort. Nevertheless, the reproducibility of the results at multiple levels (PFS, OS, ORR, immune subtyping) in two independent cohorts containing data from nearly 600 immunotherapy-treated NSCLC patients, indicate the robustness of the model. A second limitation is the relatively small sample size of patients with *KEAP1* mutation data, particularly in the SU2C study. Thus, our study was not powered to explore the impact of *KEAP1* in individual cohorts. However, the reproducibility of the data across all the clinical and molecular endpoints (e.g., immune subtyping) in the pooled SU2C/OAK/POPLAR cohort indicated the consistency of our results. Lastly, we were unable to investigate the impact of other driver mutations, particularly *NFE2L2*. The low mutational frequency of *NFE2L2* in non-squamous NSCLC, which represented most of our population, and the lack of evidence connecting *NFE2L2* mutations to reduced immunotherapy efficacy, mitigate this issue. To a similar extent, lack of data on *KRAS* mutations in the OAK/POPLAR cohort hindered subgroup analyses in non-squamous tumors, considering the relationship between *KEAP1* and *KRAS* mutations^13^.

In conclusion, results from this study indicate that KEAPness pervasively occurs across NSCLC with a different repertoire of driver mutations, and predicts survival outcomes and immune exclusion. Our data provide solid ground to the concept that transcriptomics, and integrated transcriptomics-mutational data, can significantly foster the identification of biomarkers predicting immunotherapy efficacy.

## Methods

### Cohorts and patients

The TCGA cohort, containing data from 9,229 tumor samples, was used for the identification of NRF2 target genes in NSCLC and for the pan-cancer analyses (https://xenabrowser.net).

The SU2C identification cohort contains data from NSCLC patients treated with immunotherapy. For this study, we selected those with available RNA-Seq data (n=153). Among them, 61 had matched WES. Survival outcomes, namely, PFS, OS, and ORR, were available for 136, 143, and 140 patients, respectively. Detailed information on sequencing methods are available in the original publication ^19^. Baseline clinical features included patient age at diagnosis, sex, histology, stage at diagnosis, treatment type, line of therapy, prior use of TKIs, and smoking status. PD-L1 status, assessed by the tumor proportion score (TPS), was available for 71 patients.

The OAK/POPLAR validation cohort consists of 891 patients enrolled into the phase II POPLAR trial and the phase III OAK trial, both comparing second-line atezolizumab (1200 mg IV every 3 weeks until disease progression) versus docetaxel (75 mg/m2 IV every 3 weeks) monotherapy. The study protocols and sequencing methods are detailed in the original publications ^20–22^. In the available datasets, 439 patients were treated with atezolizumab, whereas 452 patients received docetaxel chemotherapy. Regarding clinical data, histology (squamous versus non-squamous) and sex were the only clinical features reported in the datasets. Data on *KEAP1* mutations were available from 517 patients, of whom 251 treated with atezolizumab and 266 treated with docetaxel. Given that histology was reported as squamous and non-squamous in the OAK/POPLAR cohort, this definition was also applied to the SU2C and TRACERx 421 cohorts.

Genomic correlates of KEAPness were investigated in the TRACERx 421 cohort, containing matched RNA-seq and WES data from 347 non-metastatic NSCLC patients for a total of 947 tumor regions. Methods of data collection and sequencing are extensively described in the related publications ^23, 27^.

### Statistical analyses

Time to event endpoints (PFS and OS) were estimated using the Kaplan-Meier product-limit method, using the log-rank test for subgroups comparison. Multivariable Cox regression models for PFS and OS were performed by taking into account all the relevant patient-and treatment-related features reported in the original datasets. The related estimates were reported as hazard ratio (HR) and 95% confidence interval (CI). Differences in median values were estimated with the Wilcoxon test when two groups were compared. When we investigated differences among three molecular subgroups, we used the Kruskal-Wallis test followed by the Dunn’s test for pairwise comparisons, and the Benjamini-Hochberg method to control the False Discovery Rate (FDR). The relationship between categorical variables was assessed with the Pearson Chi-square test of independence. The Pearson’s correlation test was used to assess the connection among the twelve genes included in the KEAPness gene set. SPSS v21 and R (“survival” and “survminer” packages) were used for statistical analyses. The level of significance was defined as p<0.05.

### Calculation of the KEAPness signature score

The KEAPness gene set score was defined by the mean expression value of the 12 genes identified upon differential gene expression and unsupervised hierarchical clustering in the TCGA NSCLC study (*AKR1C1*, *AKR1C2*, *AKR1C3*, *AKR1B10*, *AKR1B15*, *ALDH3A1*, *CYP4F3*, *CYP4F11*, *GPX2*, *PPP2R2C*, *UGT1A9*, *UGT1A6*). In the TCGA, KEAPness was defined using a z-score ≥ 0.5, calculated with the mean absolute deviation. In order to identify a KEAPness cut-off for survival analyses, log2(TPM + 1) RNA-seq data were used for both clinical cohorts (SU2C and OAK/POPLAR) and the TRACERx 421. The optimal cut-point in predicting no durable clinical benefit (PFS<6 months) was identified with the “cutpointr” R package, using the receiver operator characteristic (ROC) curve and optimizing the Youden index via bootstrap resampling (n=2,000). For this purpose, we used a subset of patients included in the SU2C cohort, namely, non-squamous NSCLC patients who did not receive prior TKIs (n=93). The same cutoff (≥ 2.1678 versus <2.1678) was adopted for all the analyses in the SU2C, OAK/POPLAR, pooled SU2C/OAK/POPLAR, and TRACERx 421. Uncommon transcriptionally silent *KEAP1* mutation (i.e. those without enrichment for the KEAPness gene set) were classified as KEAPness-free.

### Bioinformatic analyses

For differential gene expression, carried out in the TCGA NSCLC study with the goal of identifying differentially expressed gene between *KEAP1*-*NFE2L2*-mutant and wild-type samples, we used DESeq2 with the following parameters: Log2FC ≥ 1.2 and Log2FC ≤ -1.2; padj ≤ 0.05. Unsupervised hierarchical clustering was performed with “ComplexHeatmap”, using the following settings: clustering_method_columns = “euclidean” (ward.D), column_km (k-means) = 4, row_split = 15. The study of cancer-associated pathway signatures was performed with PROGENy (Pathway RespOnsive GENes; https://saezlab.github.io/progeny/), a method that infers the activity of cancer pathways from perturbation experiments ^24^, whereas the activity of selected transcription factors was investigated with DoRothEA (https://saezlab.github.io/dorothea/) ^25^. For immune subtyping, we exploited a method that generates four subtypes (immune-depleted and non-fibrotic, immune-depleted and fibrotic, immune-enriched and non-fibrotic, immune-enriched and fibrotic) on the basis of 29 microenvironment-related signatures estimated from RNA-Seq (http://science.bostongene.com/tumor-portrait; http://science.bostongene.com/tumor-portrait) ^26^. In the TRACERx 421, the dN/dS method was used to detect positive selection in established cancer genes ^28^. The dndscv function from the dNdScv R package was run on genes from the COSMIC Cancer Gene Census (v. 98, https://cancer.sanger.ac.uk/census). Only genes with global q-values < .05 and evidence of positive selection (i.e. total number of non-synonymous mutations ≥ total number of synonymous mutations) were classified as significant. Evolutionary clustering was performed with REVOLVER, a method that exploits transfer learning to detect repeated evolution ^29^. T cell infiltration was estimated from WES data using the T cell ExTRECT method ^30^.

## Data availability

WES and RNA-Seq data related to the TCGA studies are freely available at https://xenabrowser.net. Details regarding data availability of the SU2C cohort are provided in the original publication ^19^. For this study, an additional patient treated at the Dana-Farber Cancer Institute (Boston, MA, USA) was included. Data related to the OAK/POPLAR trials are deposited to the European Genome-phenome Archive (EGA) (Study ID EGAS00001005013. Datasets: EGAD00001007703, EGAD00001008550, EGAD00001008390, EGAD00001008391, EGAD00001008548, EGAD00001008549). Data access was granted by Genentech to Dr. M. Maugeri-Saccà for the specific purpose of this study (Project Title: Mapping NRF2 activity in non-small cell lung cancer treated with immune checkpoint inhibitors). TRACERx 421 data are available from the original publication ^23^.

## Data Availability

All data produced in the present work are contained in the manuscript

## Acknowledgements

M. Maugeri-Saccà is supported by the Italian Association for Cancer Research (AIRC) under MFAG 2019 - project ID. 22940, and the Italian Ministry of Health (MoH)-project ID. GR-2016-02362025. N.M. is a Sir Henry Dale Fellow, jointly funded by the Wellcome Trust and the Royal Society (Grant Number 211179/Z/18/Z), and also receives funding from Cancer Research UK Lung Cancer Centre of Excellence, Rosetrees, and the NIHR BRC at University College London Hospitals. B. Ricciuti is supported by The Conquer Cancer Foundation of the American Society for Clinical Oncology and the Society for Immunotherapy of Cancer. G. Caravagna is supported by the Italian Association for Cancer Research (AIRC) under MFAG 2020-ID. 24913. D. Marinelli is a fellow of the PhD Network Oncology and Precision Medicine, Department of Experimental Medicine, Sapienza University of Rome. The research leading to these results has received funding from the European Union - NextGenerationEU through the Italian Ministry of University and Research under PNRR - M4C2-I1.3 Project PE_00000019 “HEAL ITALIA” to M. Maugeri-Saccà and G. Ciliberto, CUP H83C22000550006. The views and opinions expressed are those of the authors only and do not necessarily reflect those of the European Union or the European Commission. Neither the European Union nor the European Commission can be held responsible for them.

## Extended data figures

**Extended Data** Figure 1**. KEAPness and pathway-level signatures (PROGENy) in NSCLC (TCGA).** Box plots illustrating the relationship between PROGENy signatures and KEAPness (androgen, EGFR, estrogen, hypoxia, MAPK, PI3K, TGFb, TNFa, TRAIL, VEGF, and WNT). JAK-STAT, p53 and NFkB are reported in figure 2.

**Extended Data** Figure 2**. KEAPness and the tumor immune microenvironment (TIME) in the TCGA NSCLC study. a,** Heatmap illustrating the 29 TIME signatures exploited for generating four immune subtypes (BostonGene) in relation to the KEAPness state.

**Extended Data** Figure 3**. Optimal cutoff of the KEAPness signature. a, b** ROC-AUC curve and bootstrap resampling to identify the optimal KEAPness cutoff in predicating lack of clinical benefit in a subset of patients included in the SU2C identification cohort (n=93, lung adenocarcinoma patients who did not receive prior tyrosine kinase inhibitors). Lack of durable clinical benefit was defined as a progression-free survival shorter than 6 months.

**Extended Data** Figure 4**. Multivariable Cox regression models in the SU2C and OAK/POPLAR cohorts. a-d,** Forest plots illustrating multivariable Cox regression models for PFS and OS in the SU2C (a, b) and OAK/POPLAR (c, d) cohorts.

**Extended Data** Figure 5**. KEAPness and pathway-level signatures (PROGENy) in the pooled SU2C/OAK/POPLAR cohort.** Box plots illustrating the relationship between 14 PROGENy signatures and KEAPness, *KEAP1*-mutant, and KEAPness-free NSCLC.

**Extended Data** Figure 6**. KEAPness transcriptional heterogeneity in relation to histology in the TRACERx 421 cohort. a, b,** Dumbbell plot showing KEAPness heterogeneity at the individual patient level in non-squamous (a) and squamous (b) tumors. **c,** Stacked bar chart illustrating the frequency of KEAPness heterogeneity according to histology.

## Supplementary figures and tables

**Supplementary Figure 1. KEAPness in lung adenocarcinoma (LUAD) and lung squamous cell carcinoma (LUSC) in the TCGA. a,** Stacked bar chart illustrating the distribution of KEAPness in relation to histology (KEAPness: KEAPness+; KEAPness-free: KEAPness-; *KEAP1*-mutant: mKEAP1).

**Supplementary Figure 2. KEAPness and cancer-associated pathway signatures at the pan-cancer level.** Box plots illustrating the relationship between KEAPness and PROGENy pathway signatures across the different cancer types (tumors that are prevalently KEAPness-free were excluded).

**Supplementary Figure 3. Immune-related processes and the KEAPness-dominant phenotype in the SU2C and OAK/POPLAR cohorts. a, b,** Heatmaps illustrating the 29 TIME signatures used for immune subtyping (BostonGene) in relation to the KEAPness state in the SU2C (a) and OAK/POPLAR (b) cohorts.

**Supplementary Figure 4. PD-L1 tumor proportion score (TPS) in the SU2C identification cohort. a,** Stacked bar chart showing the distribution of PD-L1 TPS (<50% and ≥50%) in KEAPness-dominant and KEAPness-free NSCLC.

**Supplementary Figure 5. Sensitivity analyses in the SU2C cohort upon exclusion of patients who received prior targeted agents. a, b,** Kaplan-Meier survival curves for progression-free survival (PFS) and overall survival (OS). **c, d**, Forest plots illustrating multivariate Cox regression models for PFS and OS. **e,** Stacked bar chart for overall response rate (ORR).

**Supplementary Figure 6: Impact of the KEAPness-dominant phenotype in advanced NSCLC treated with chemotherapy (docetaxel) in the OAK/POPLAR trials. a, b,** Kaplan-Meier survival curves for progression-free survival (PFS) and overall survival (OS). **c,** Stacked bar chart illustrating the differences in overall response rate (ORR) between KEAPness-dominant and KEAPness-free NSCLC.

**Supplementary Figure 7. KEAPness enrichment in relation to histology in the SU2C identification cohort and OAK/POPLAR validation cohort. a, b**, Stacked bar charts showing the difference in the distribution of the KEAPness-dominant phenotype between non-squamous and squamous tumors in the SU2C (a) and OAK/POPLAR (b) cohorts.

**Supplementary Figure 8: Impact of the KEAPness-dominant phenotype according to tumor histology in the pooled SU2C/OAK/POPLAR cohort. a, b,** Kaplan-Meier survival curves for progression-free survival (PFS) and overall survival (OS) comparing KEAPness-dominant and KEAPness-free non-squamous NSCLC. **c, d,** Kaplan-Meier survival curves for progression-free survival (PFS) and overall survival (OS) comparing the same subgroups in squamous lung cancer. **e, f,** Kaplan-Meier survival curves for progression-free survival (PFS) and overall survival (OS) comparing KEAPness-dominant squamous and non-squamous tumors. **g, h,** Kaplan-Meier survival curves for progression-free survival (PFS) and overall survival (OS) comparing KEAPness-free squamous and non-squamous tumors.

**Supplementary Table 1.**
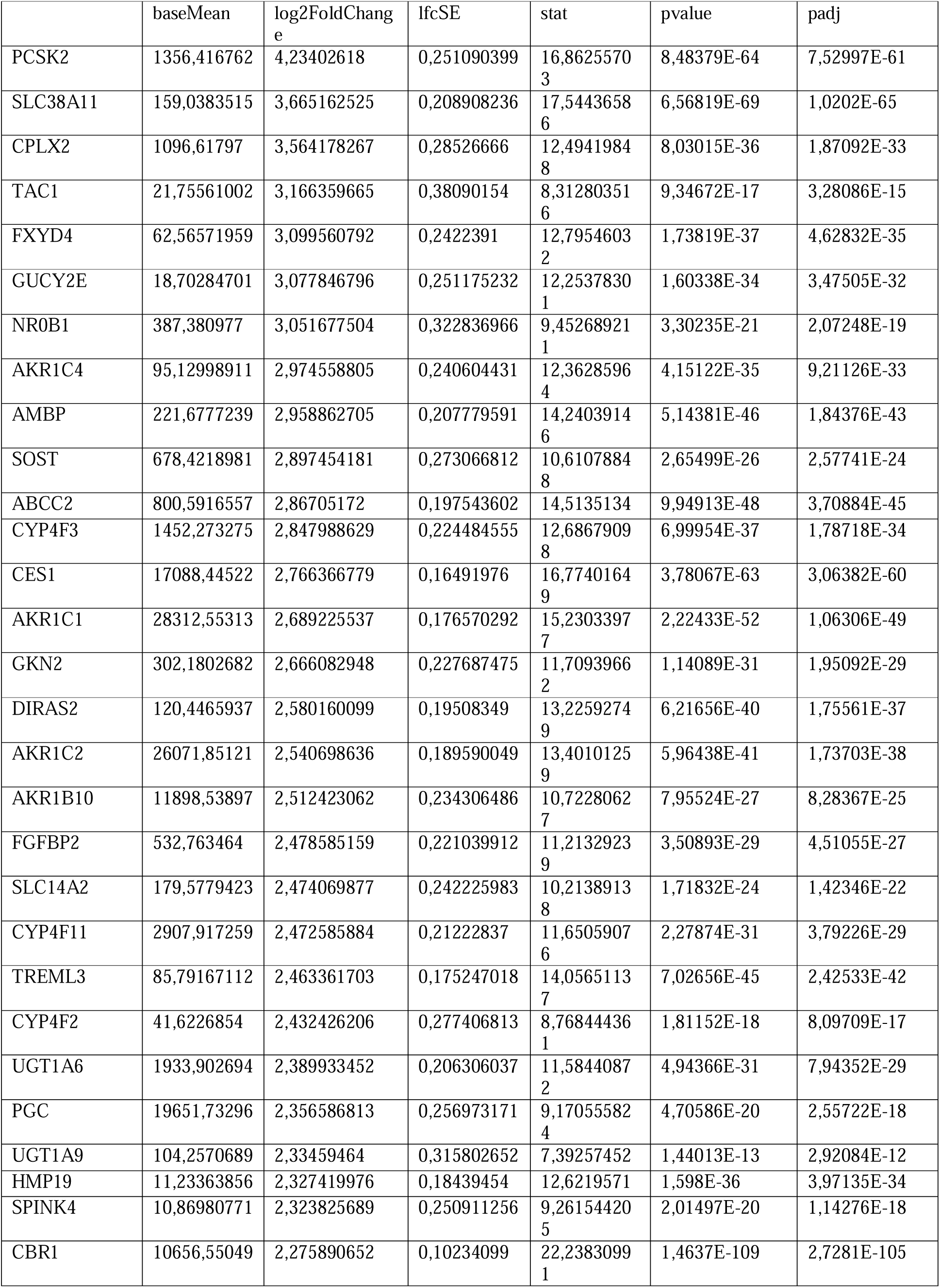
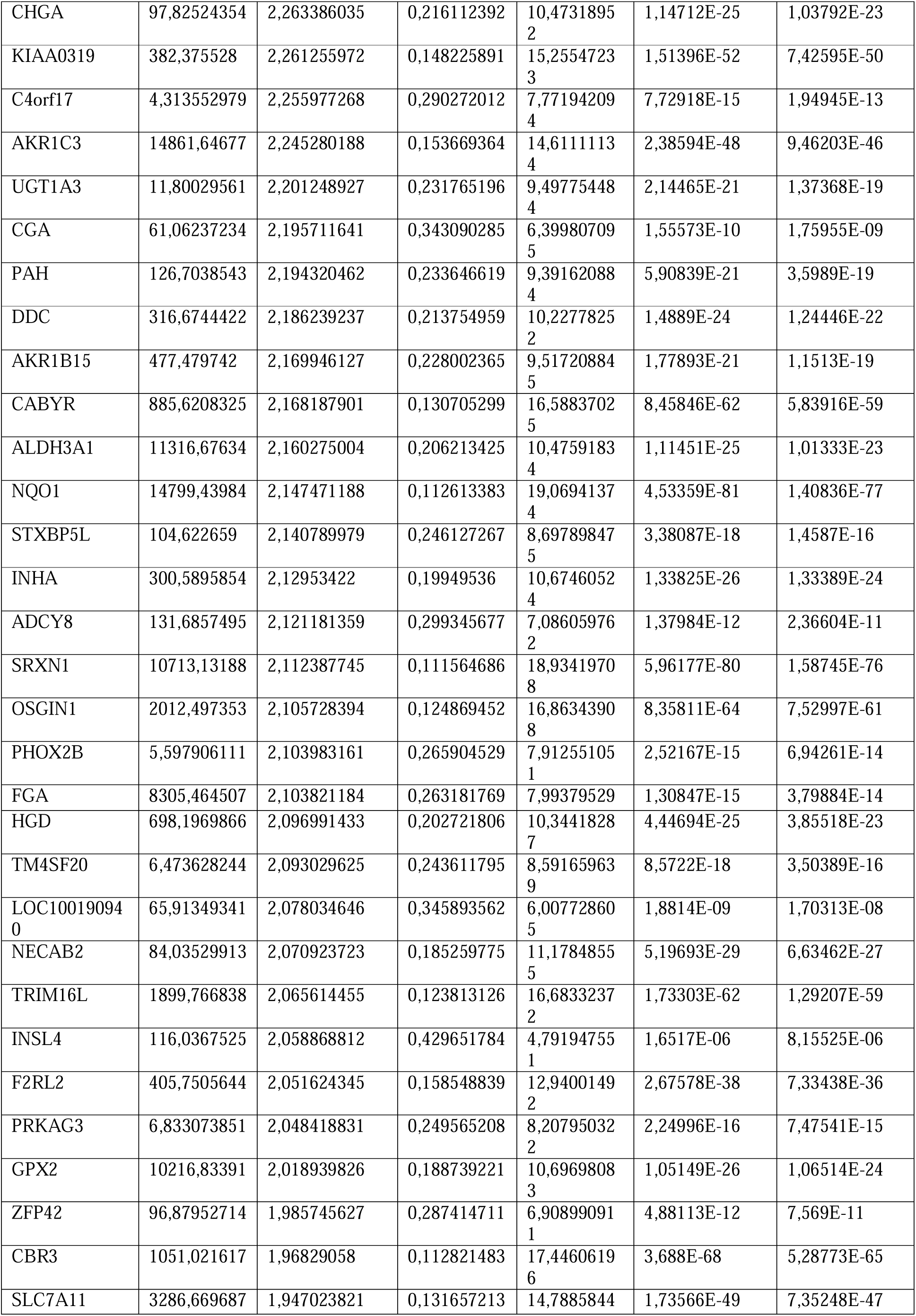
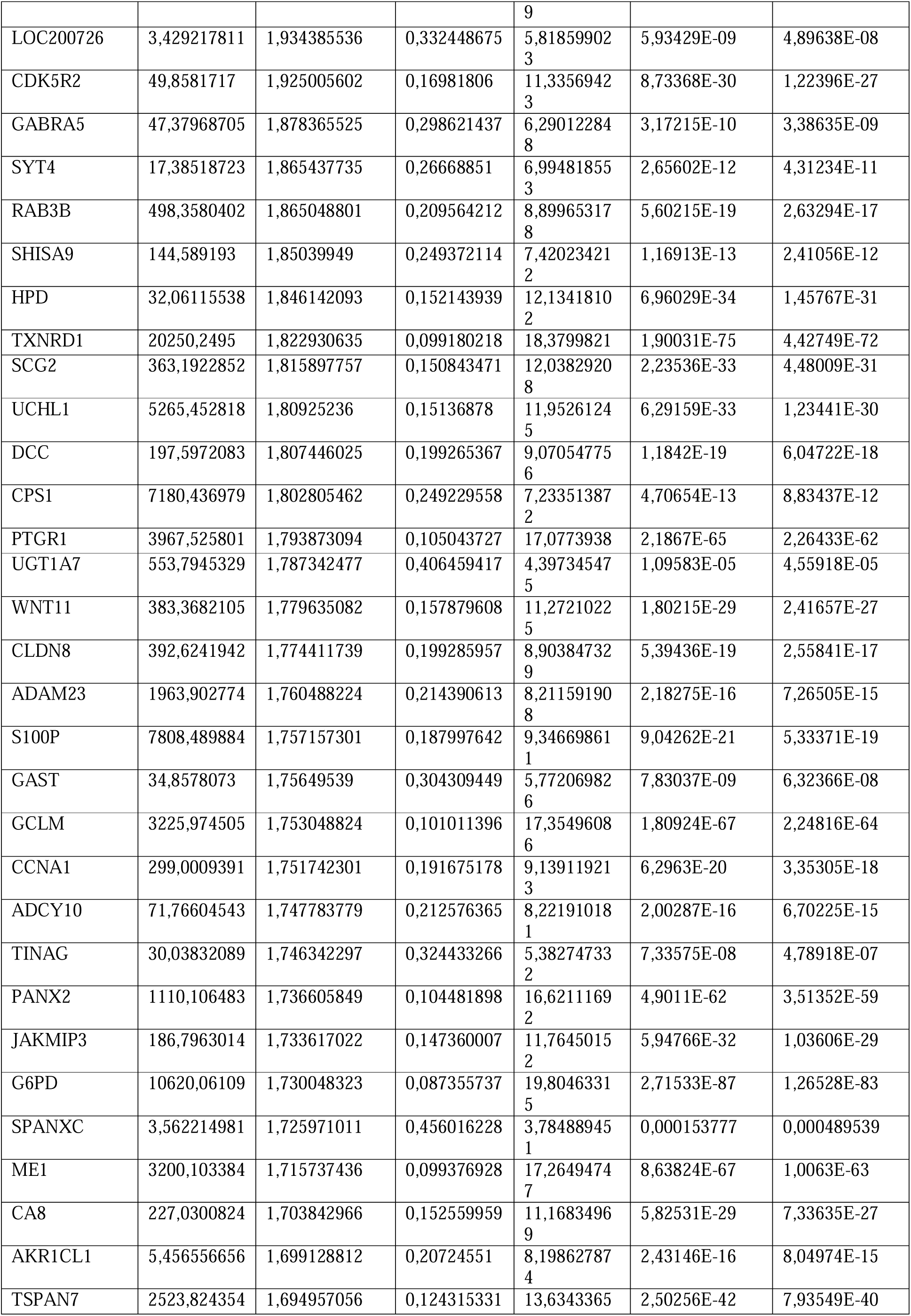
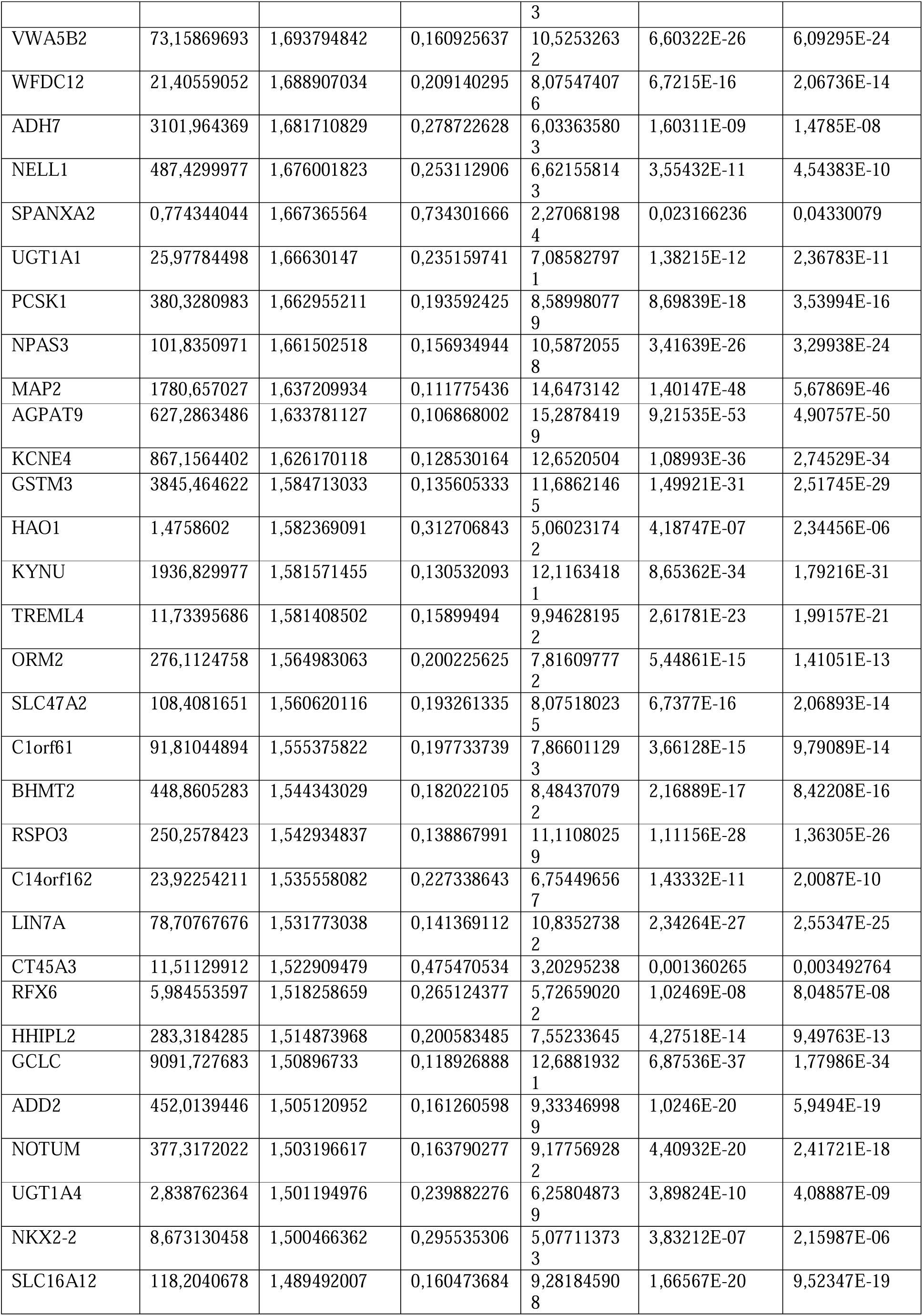
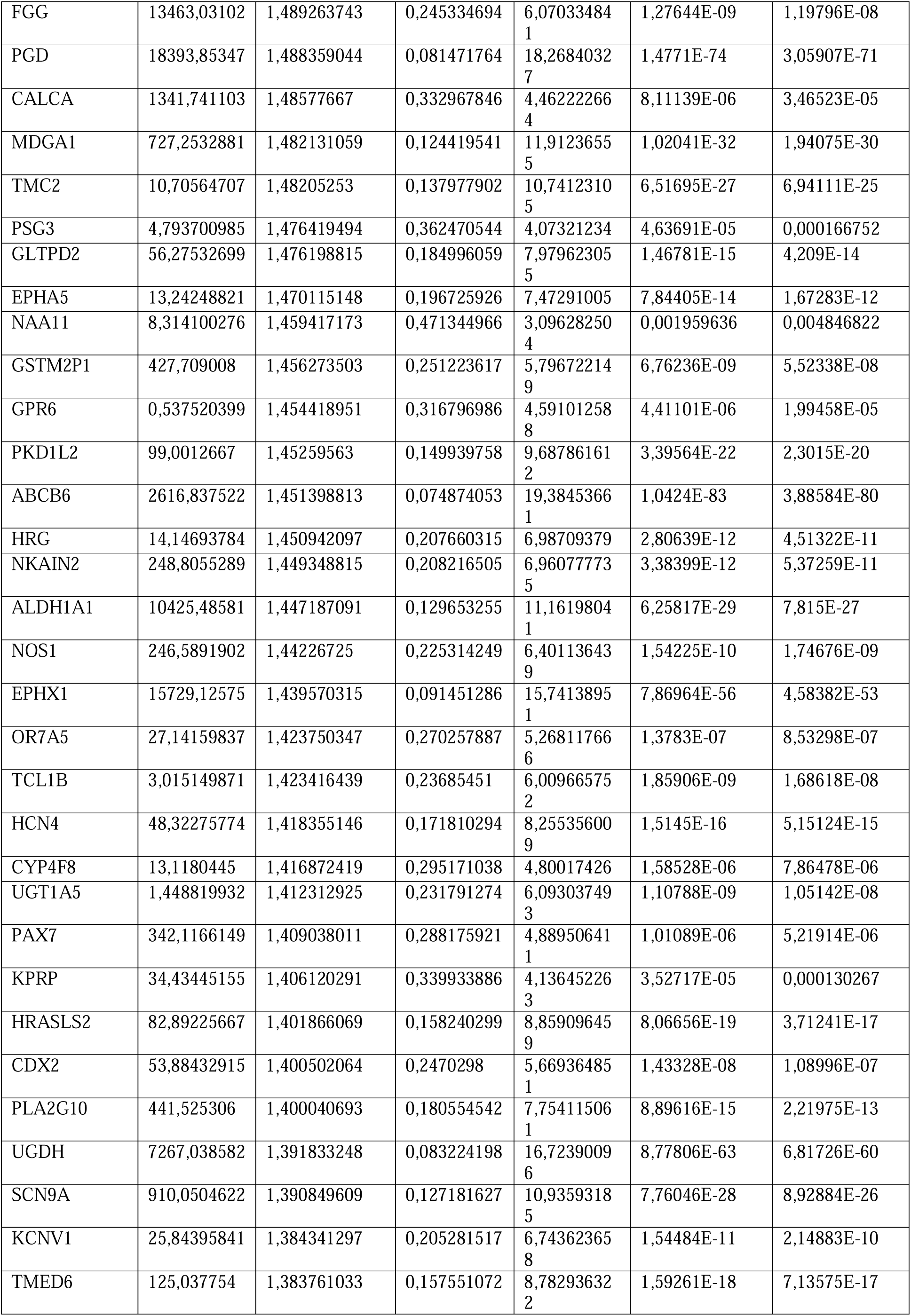
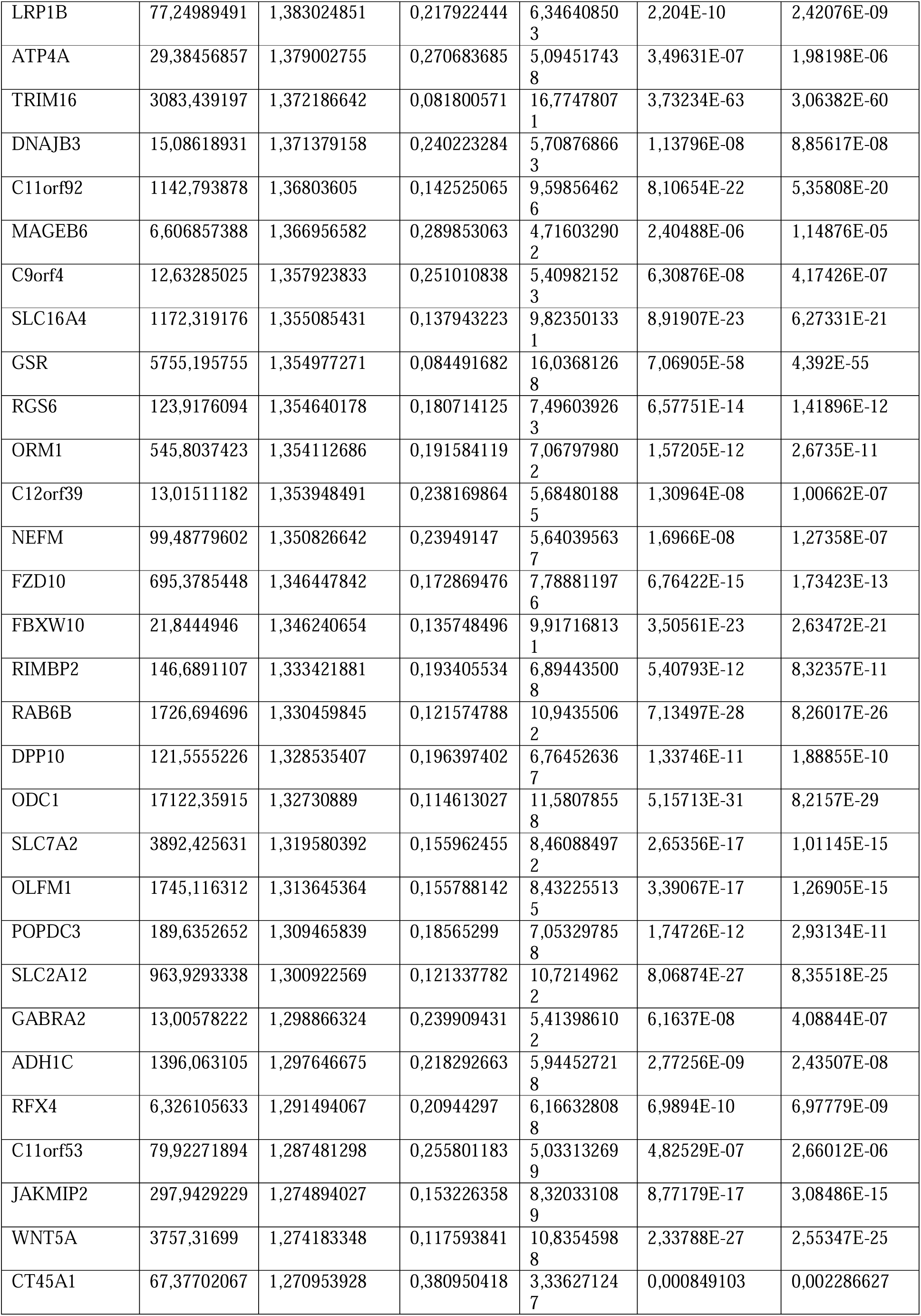
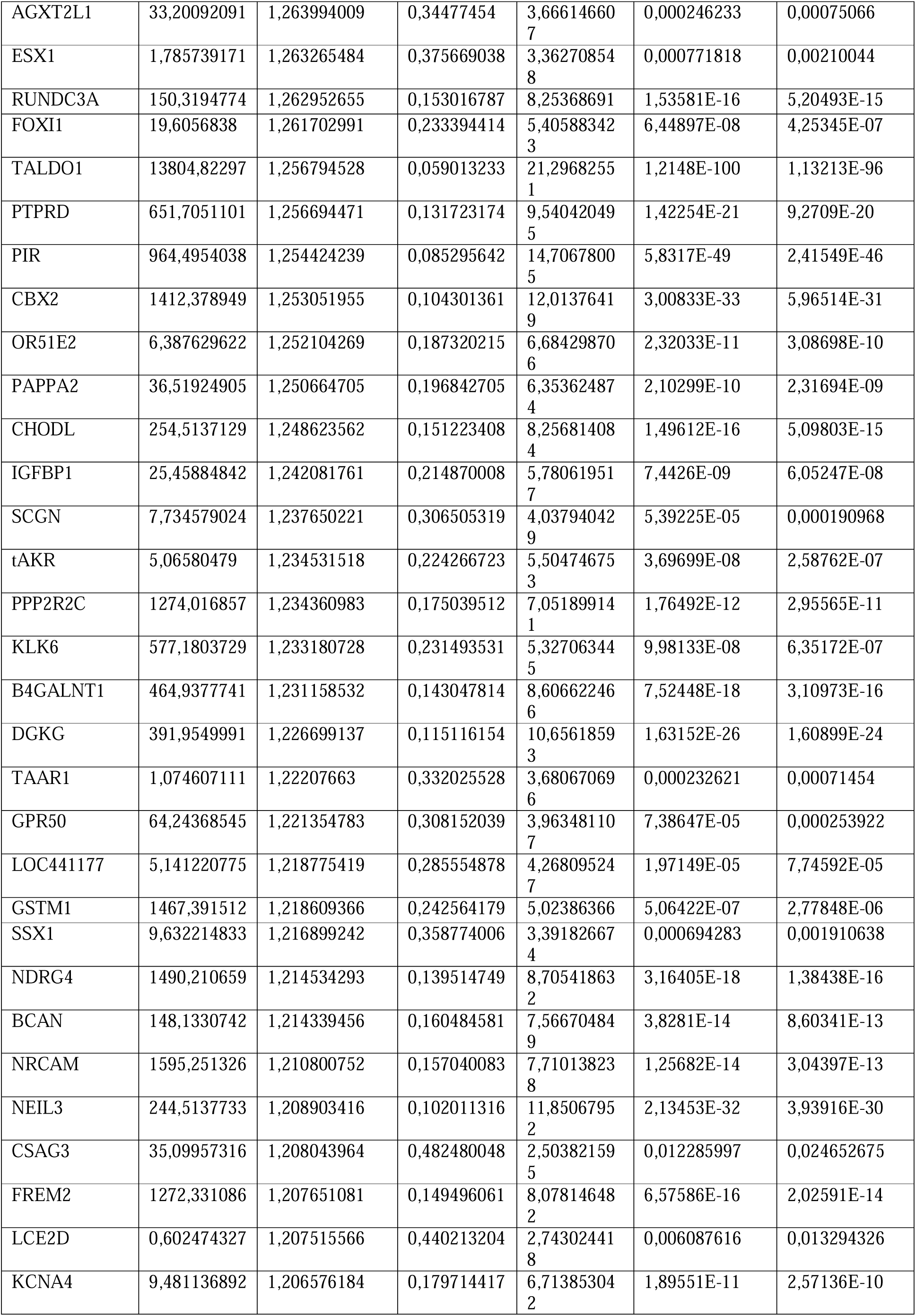
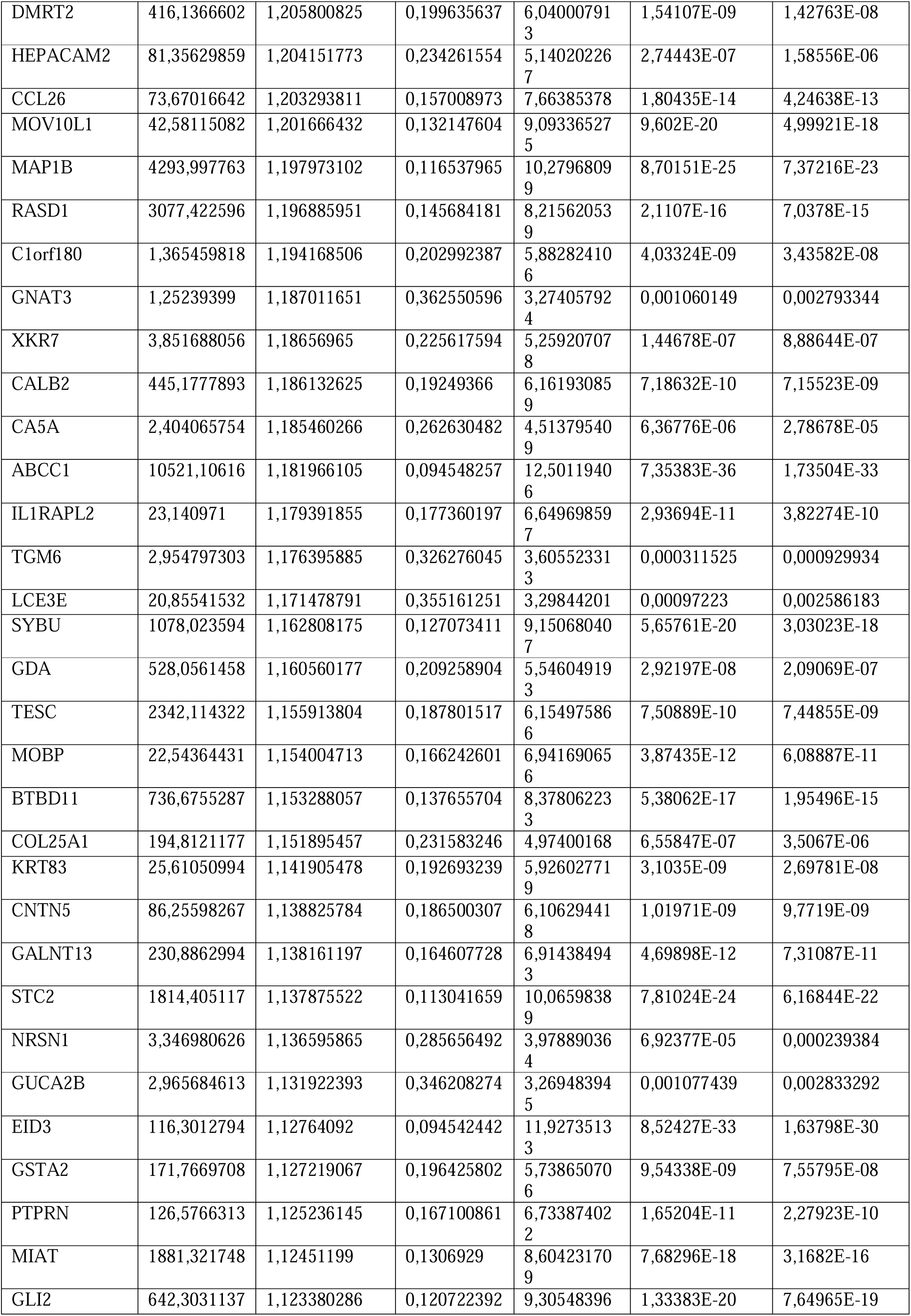
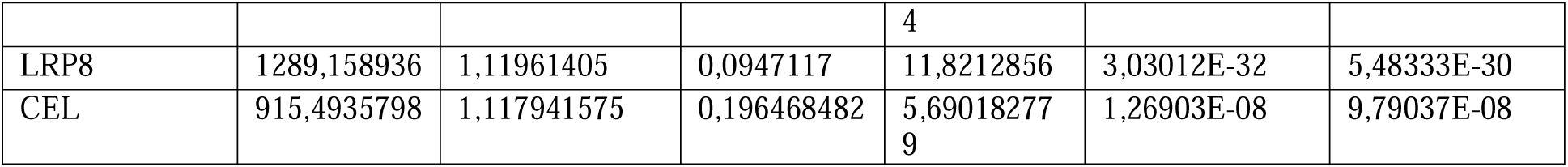
List of up-regulated genes identified by differential gene expression when comparing *KEAP1*-*NFE2L2*-mutant and wild-type NSCLC in the TCGA study.

**Supplementary Table 2.** Clinical features of the patients included in the SU2C identification cohort.

| Clinical characteristic |  | n (%) |
| --- | --- | --- |
| ***Histology*** |  |  |
|  | Non-Squamous | 117 (76.5) |
|  | Squamous | 32 (20.9) |
|  | NA | 4 (2.6) |
| ***Stage at diagnosis*** |  |  |
|  | I | 14 (9.2) |
|  | II | 10 (6.5) |
|  | III | 35 (22.9) |
|  | IV | 82 (53.6) |
|  | NA | 12 (7.8) |
| ***Sex*** |  |  |
|  | Male | 66 (43.1) |
|  | Female | 85 (55.6) |
|  | NA | 2 (1.3) |
| ***Line of treatment*** |  |  |
|  | 1 | 48 (31.4) |
|  | 2 | 64 (41.8) |
|  | 3 | 23 (15.0) |
|  | ≥ 4 | 14 (9.2) |
|  | NA | 4 (2.6) |
| ***Smoking*** |  |  |
|  | Never | 23 (15.0) |
|  | Former | 105 (68.7) |
|  | Current | 21 (13.7) |
|  | NA | 4 (2.6) |
| ***Age (years)*** |  |  |
|  | ≥ 65 | 72 (47.1) |
|  | < 65 | 77 (50.3) |
|  | NA | 4 (2.6) |
| ***Prior TKIs*** |  |  |
|  | yes | 12 (7.8) |
|  | no | 137 (89.6) |
|  | NA | 4 (2.6) |
| ***ICI treatment type*** |  |  |
|  | Monotherapy | 135 (88.2) |
|  | Combination | 18 (11.8) |

Abbreviations: NA, Not Available. TKIs, Tyrosine Kinase Inhibitors. ICI, immune checkpoint inhibitor.

**Supplementary Table 3.** Association between the KEAPness-dominant phenotype and baseline characteristics of NSCLC patients included in the SU2C identification cohort.

| Clinical characteristic | Group (n) | | $\chi^2$ test (p) |
| --- | --- | --- | --- |
|  | KEAPness-free | KEAPness-dominant |  |
| **Histology** |  |  |  |
| Non-Squamous | 63 | 54 | 0.079 |
| Squamous | 11 | 21 |  |
| **Stage at diagnosis** |  |  |  |
| Other | 28 | 31 | 0.5 |
| IV | 44 | 38 |  |
| **Sex** |  |  |  |
| Female | 41 | 42 | 1.0 |
| Male | 33 | 33 |  |
| **Line of treatment** |  |  |  |
| 1-2 | 53 | 59 | 0.4 |
| $\geq 3$ | 21 | 16 | |
| **Smoking** |  |  |  |
| Never/Former | 66 | 62 | 0.36 |
| Current | 8 | 13 |  |
| **Prior TKIs** |  |  |  |
| yes | 12 | 0 | < 0.01 |
| no | 62 | 75 |  |
| **ICI treatment type** |  |  |  |
| Monotherapy | 65 | 70 | 0.73 |
| Combination | 10 | 8 |  |

Abbreviations: TKIs, Tyrosine Kinase Inhibitors. ICI, immune checkpoint inhibitor.

**Supplementary Table 4.** Clinical features of the patients included in the OAK/POPLAR validation cohort and association with the KEAPness-dominant phenotype.

| Clinical characteristic |  | n (%) |  |  |
| --- | --- | --- | --- | --- |
| **Histology** |  |  |  |  |
|  | Non-Squamous | 629 (70.6) |  |  |
|  | Squamous | 262 (29.4) |  |  |
| **Sex** |  |  |  |  |
|  | Female | 562 (63.1) |  |  |
|  | Male | 329 (36.9) |  |  |
| **Therapy** |  |  |  |  |
|  | Atezolizumab | 439 (49.3) |  |  |
|  | Docetaxel | 452 (50.7) |  |  |
| | | Group (n) | | $\chi^2$ test (p) |
|  |  | KEAPness-free | KEAPness-dominant |  |
| **Histology** |  |  |  |  |
|  | Non-Squamous | 368 | 261 | < 0.01 |
|  | Squamous | 50 | 212 |  |
| **Sex** |  |  |  |  |
|  | Female | 184 | 145 | < 0.01 |
|  | Male | 234 | 328 |  |

