## Supplementary figures and images for "Transcriptional phenocopies of deleterious *KEAP1* mutations dictate survival outcomes in lung cancer treated with immunotherapy"

### Extended data Figure 1

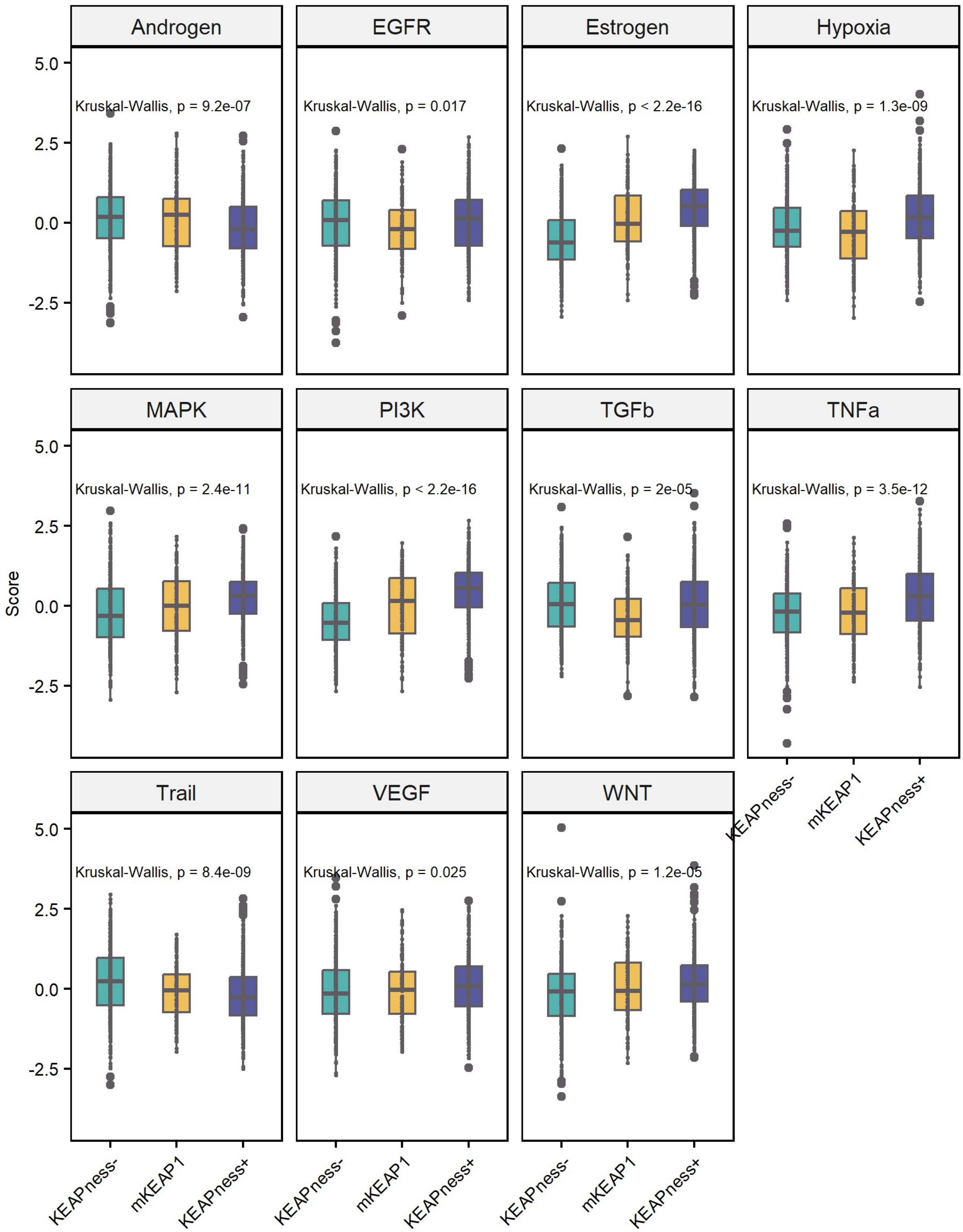

### Extended data Figure 2

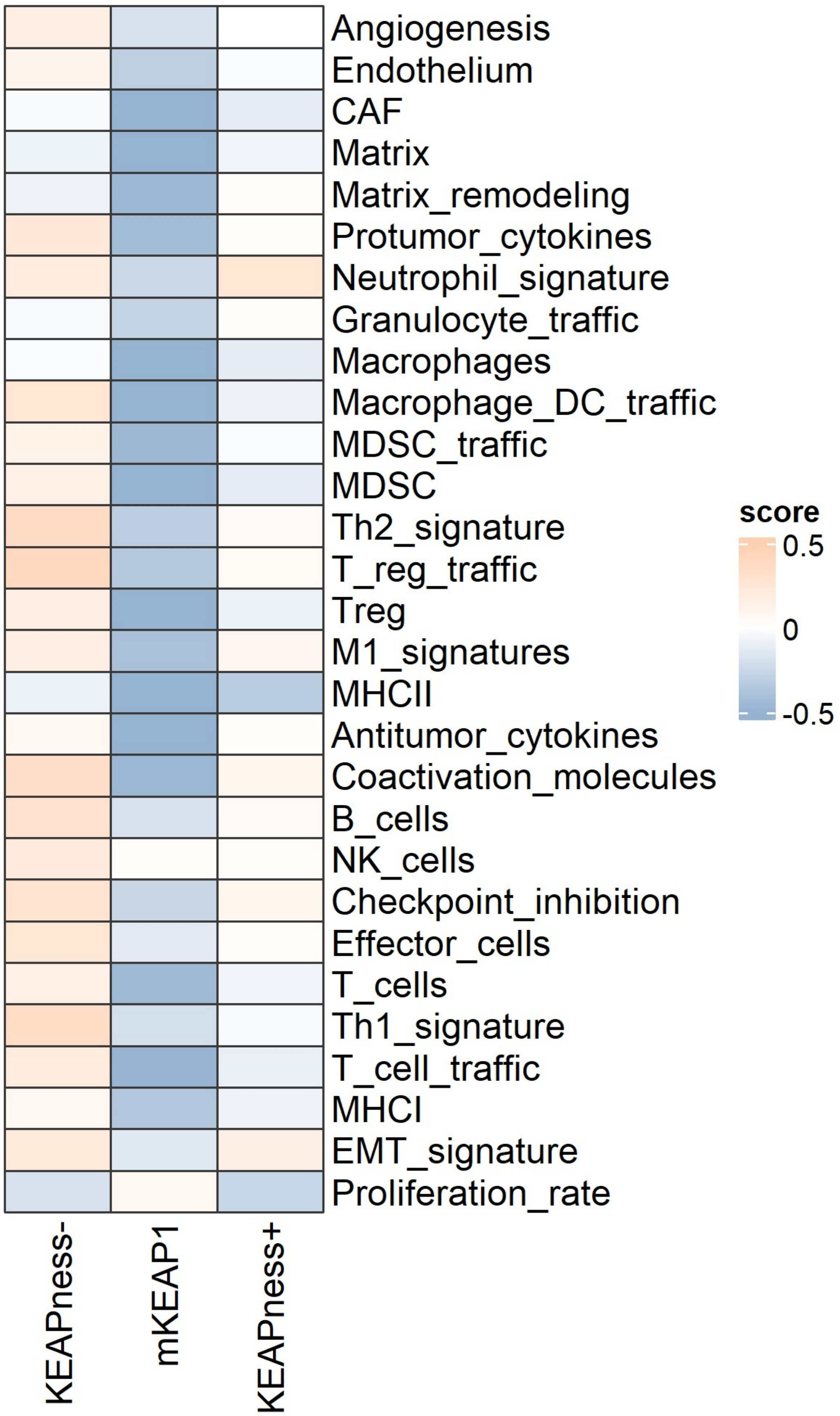

### Extended data Figure 3

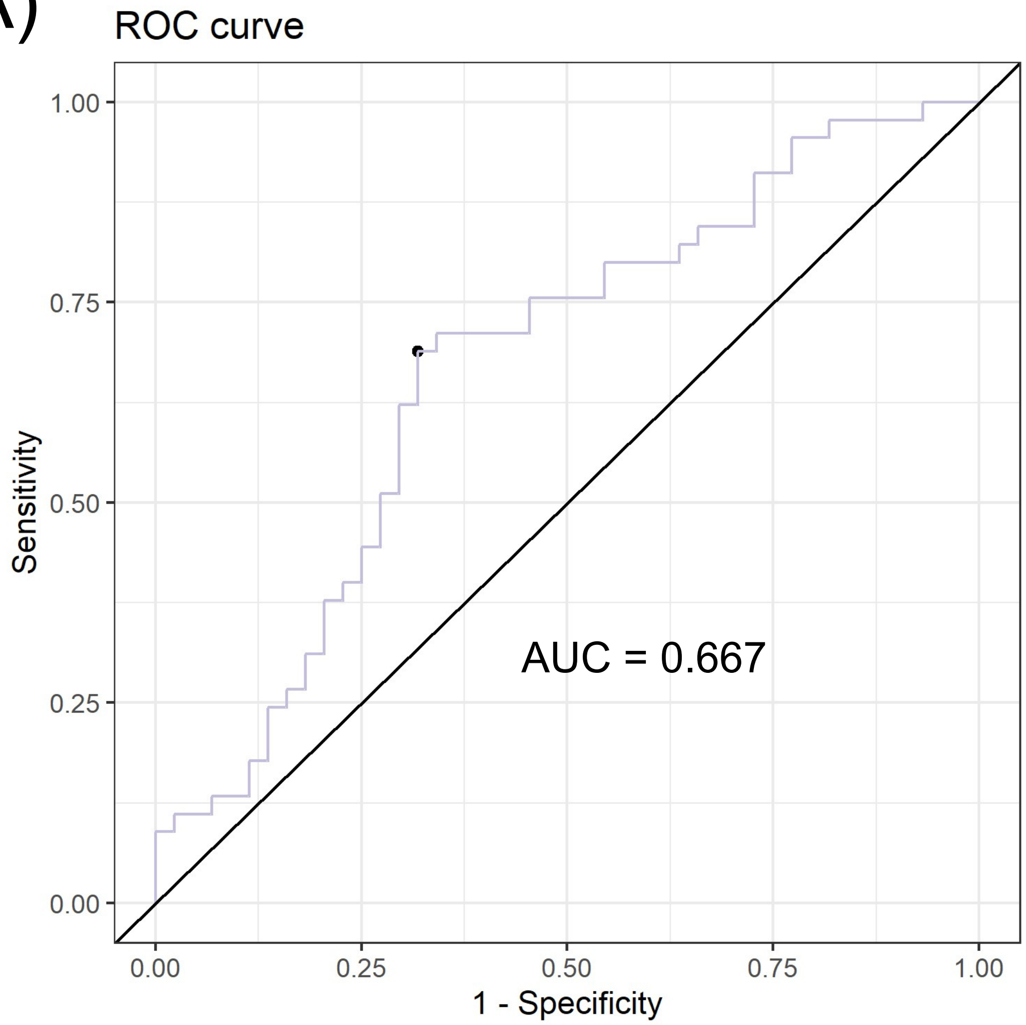

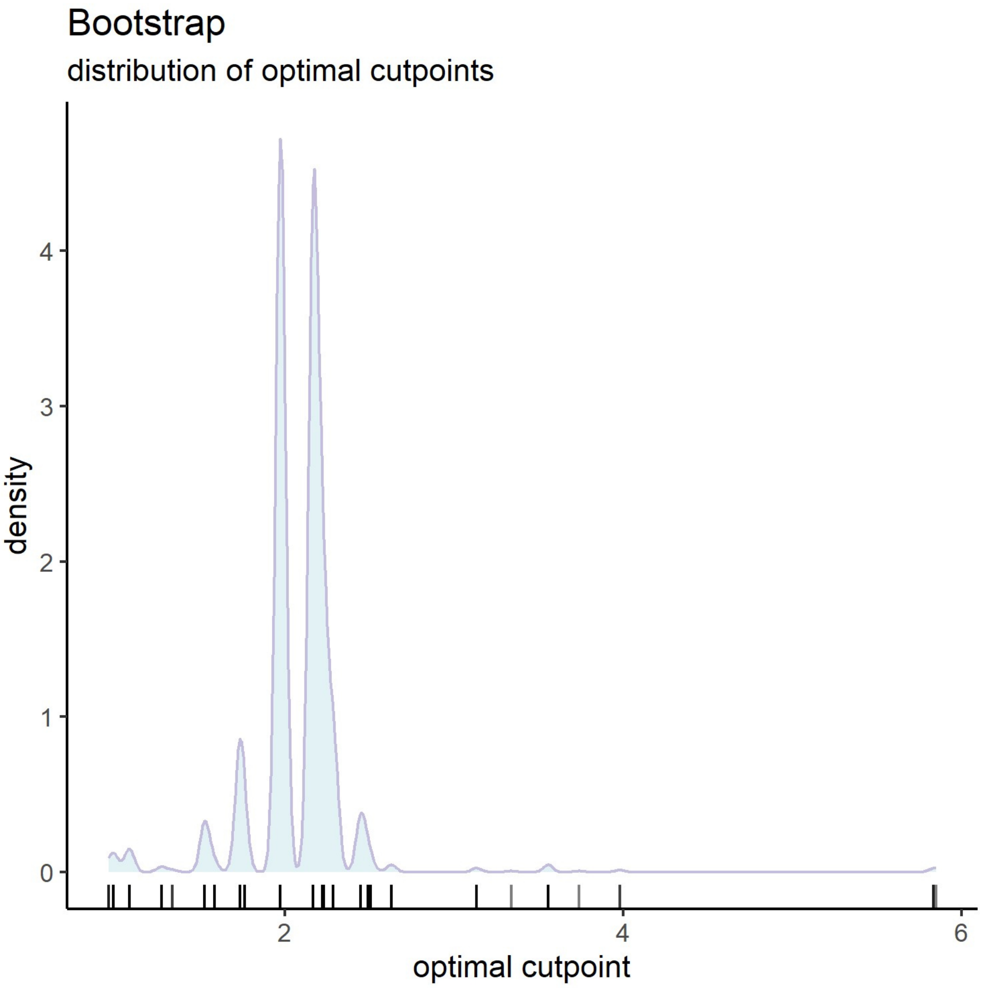

### Extended data Figure 4

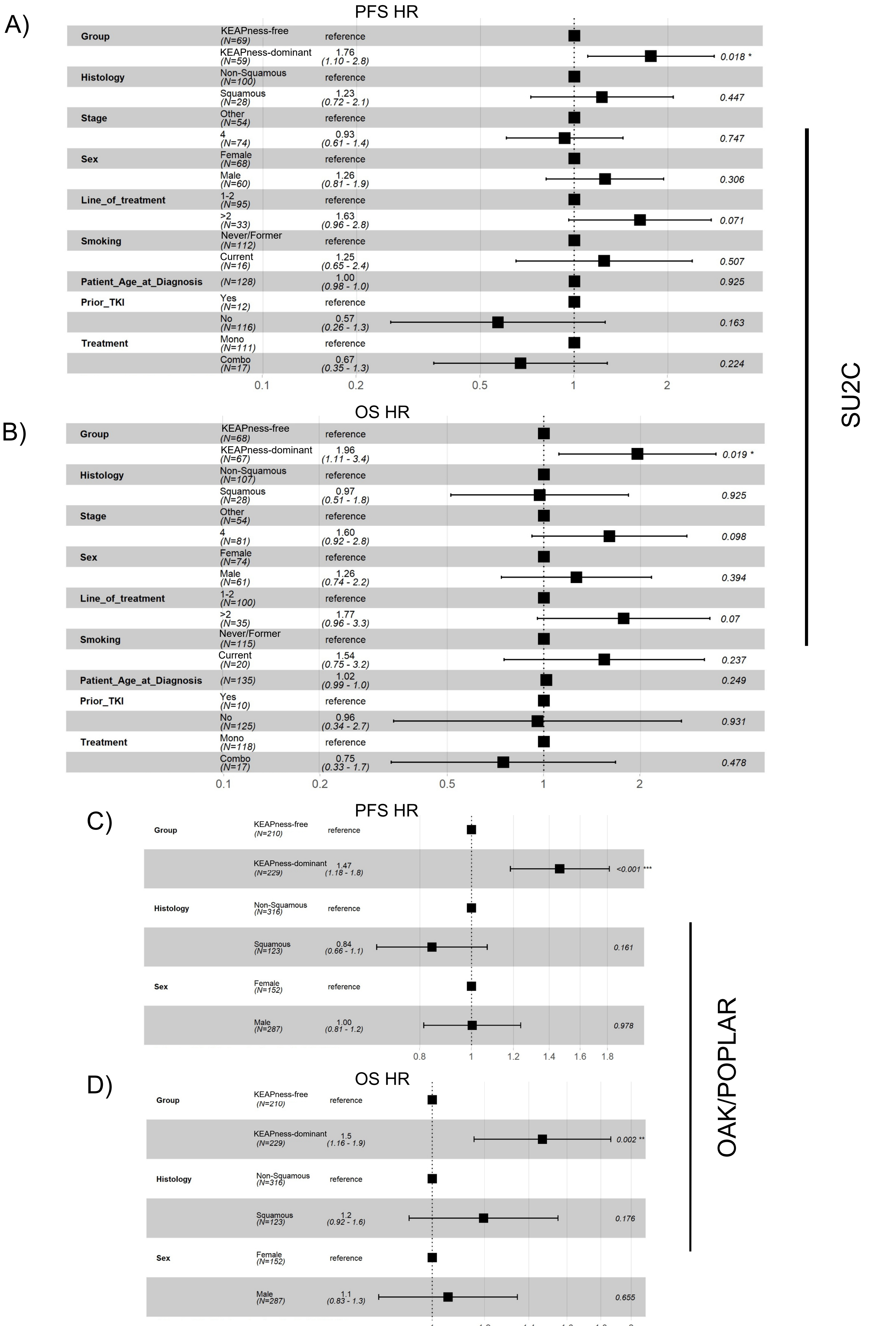

### Extended data Figure 5

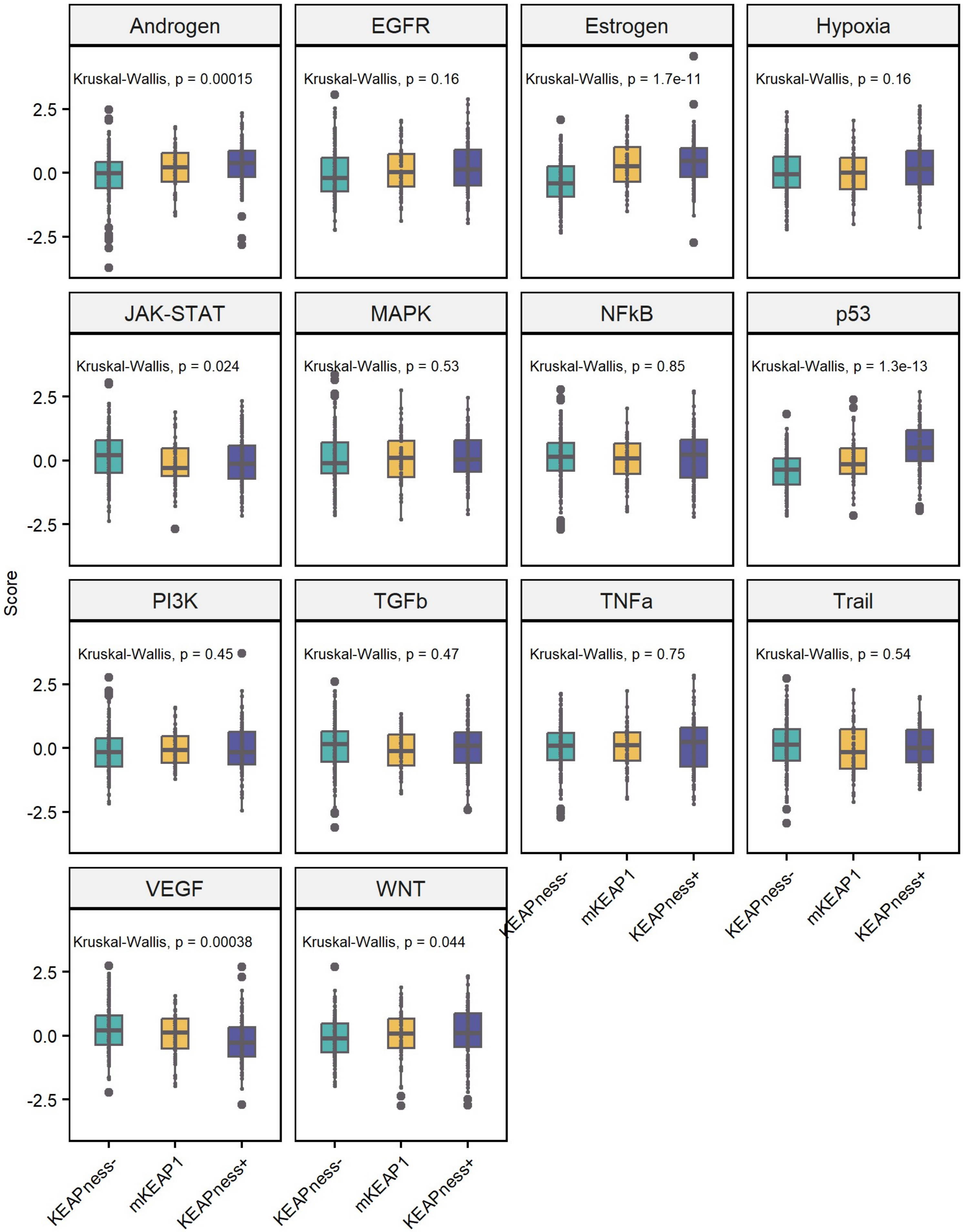

### Extended data Figure 6

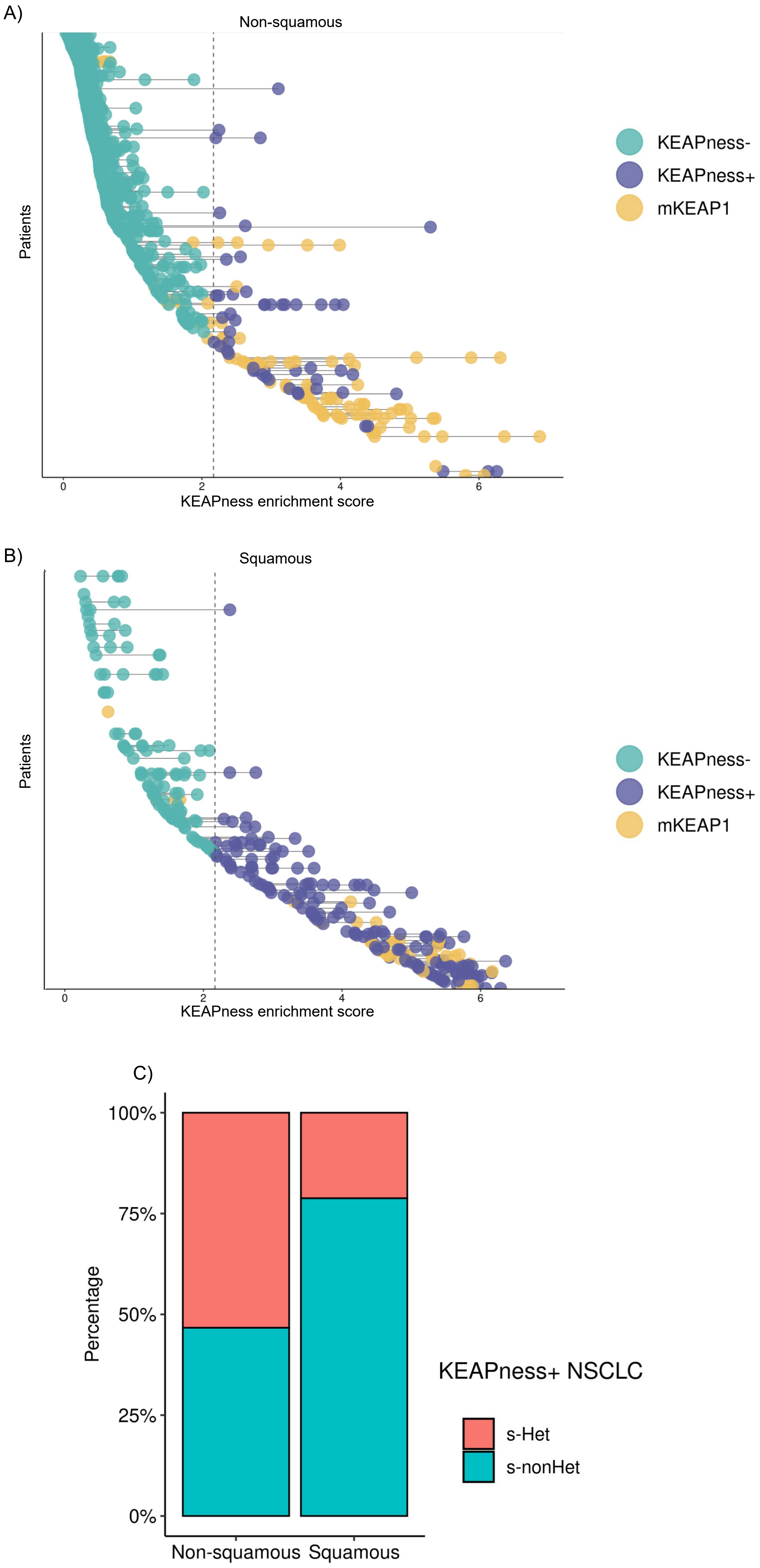

### Supplementary Figure 1

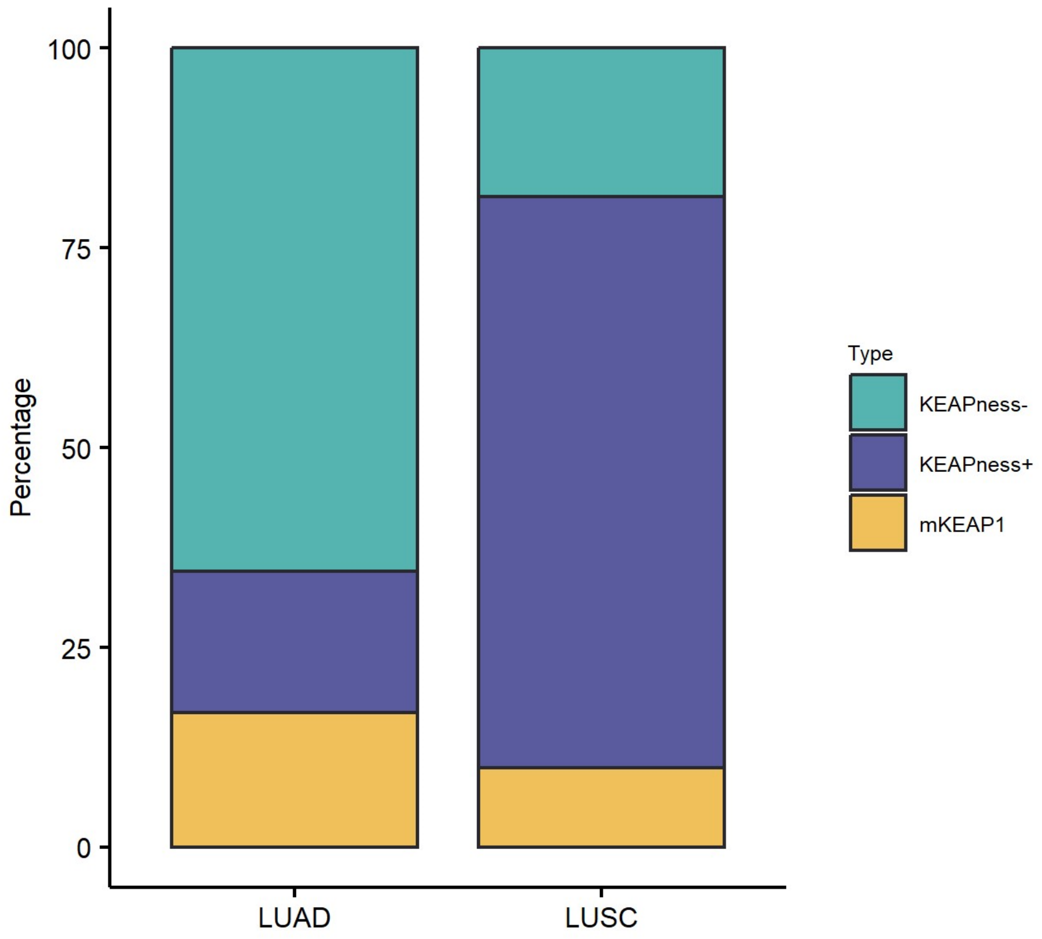

### Supplementary Figure 2

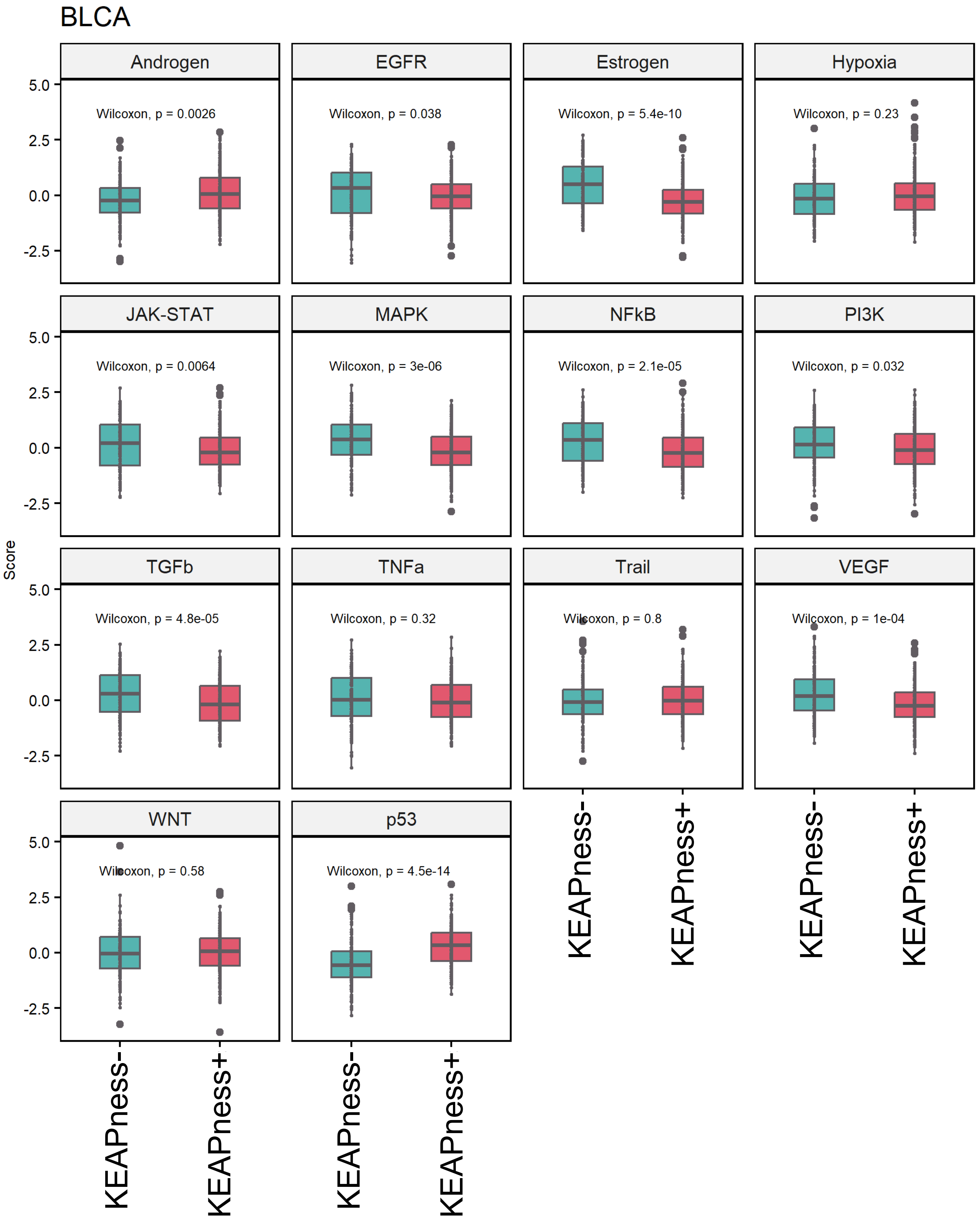

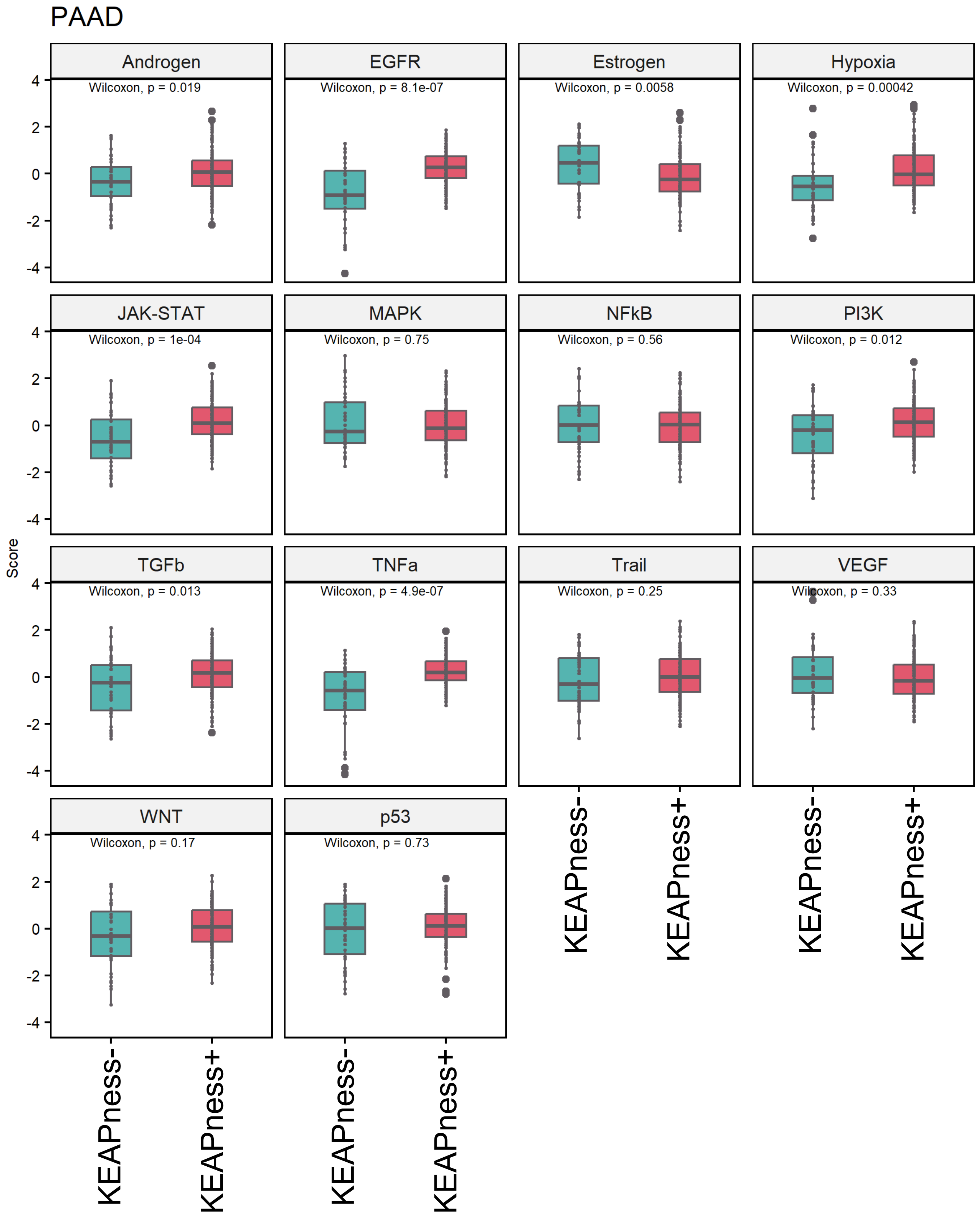

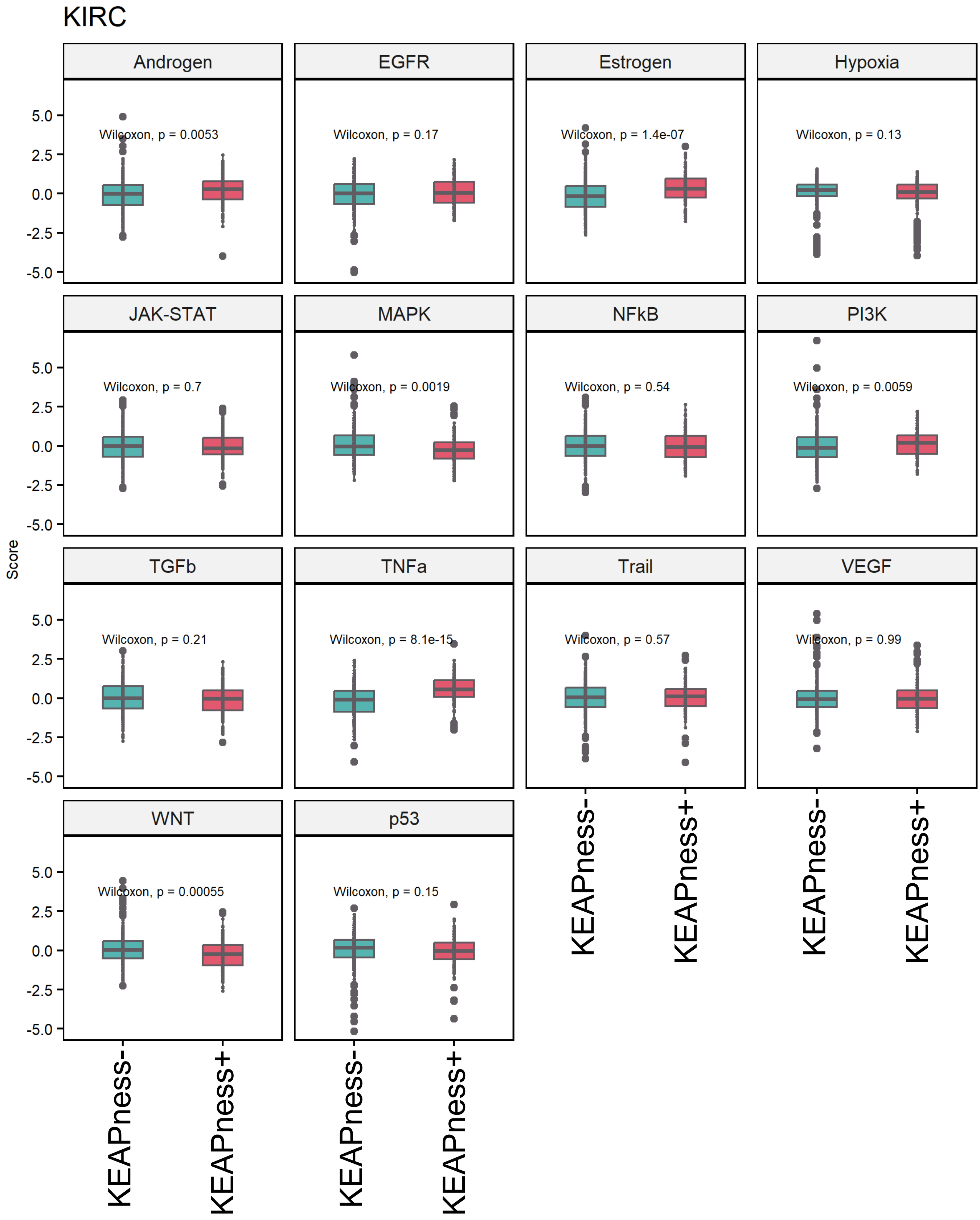

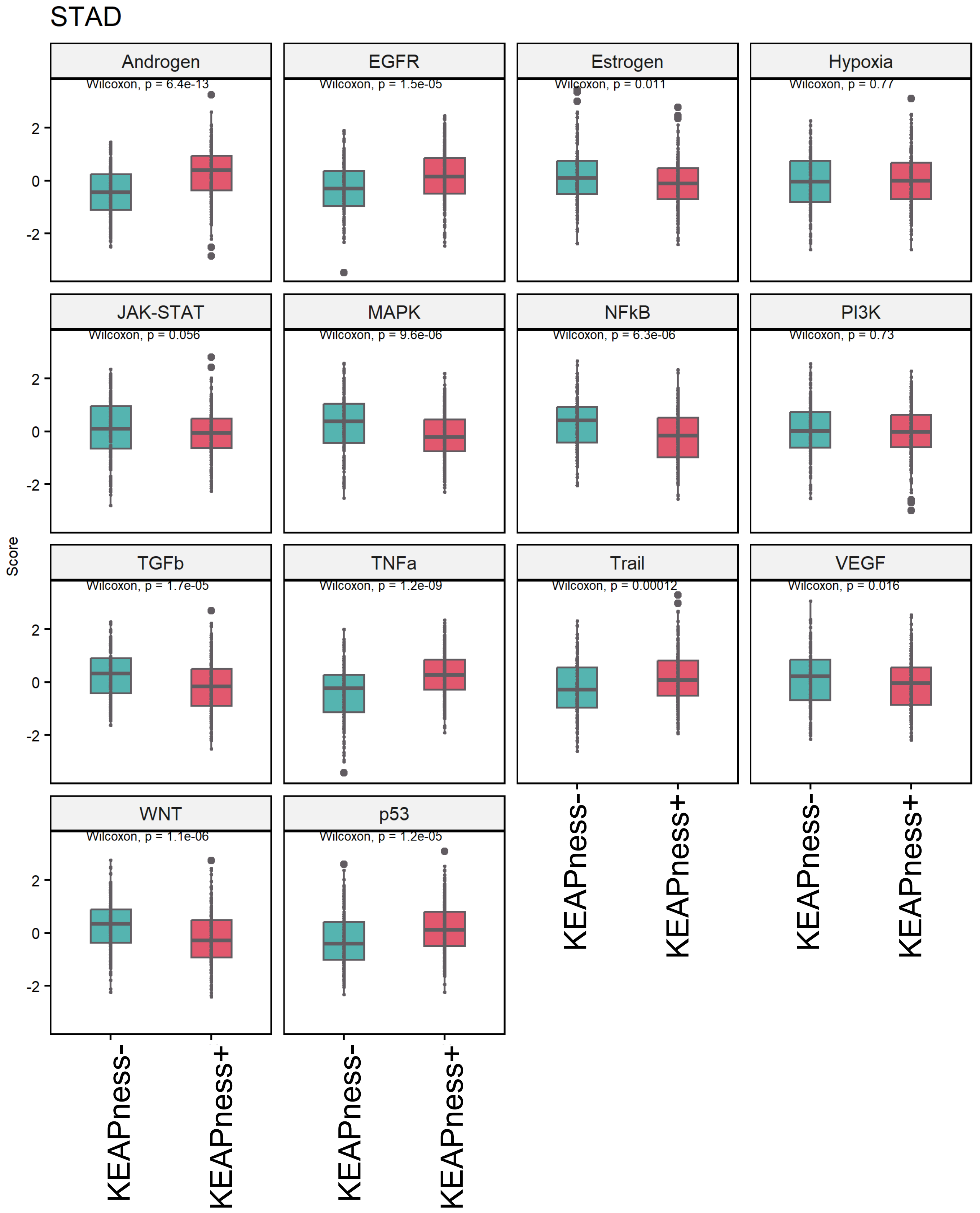

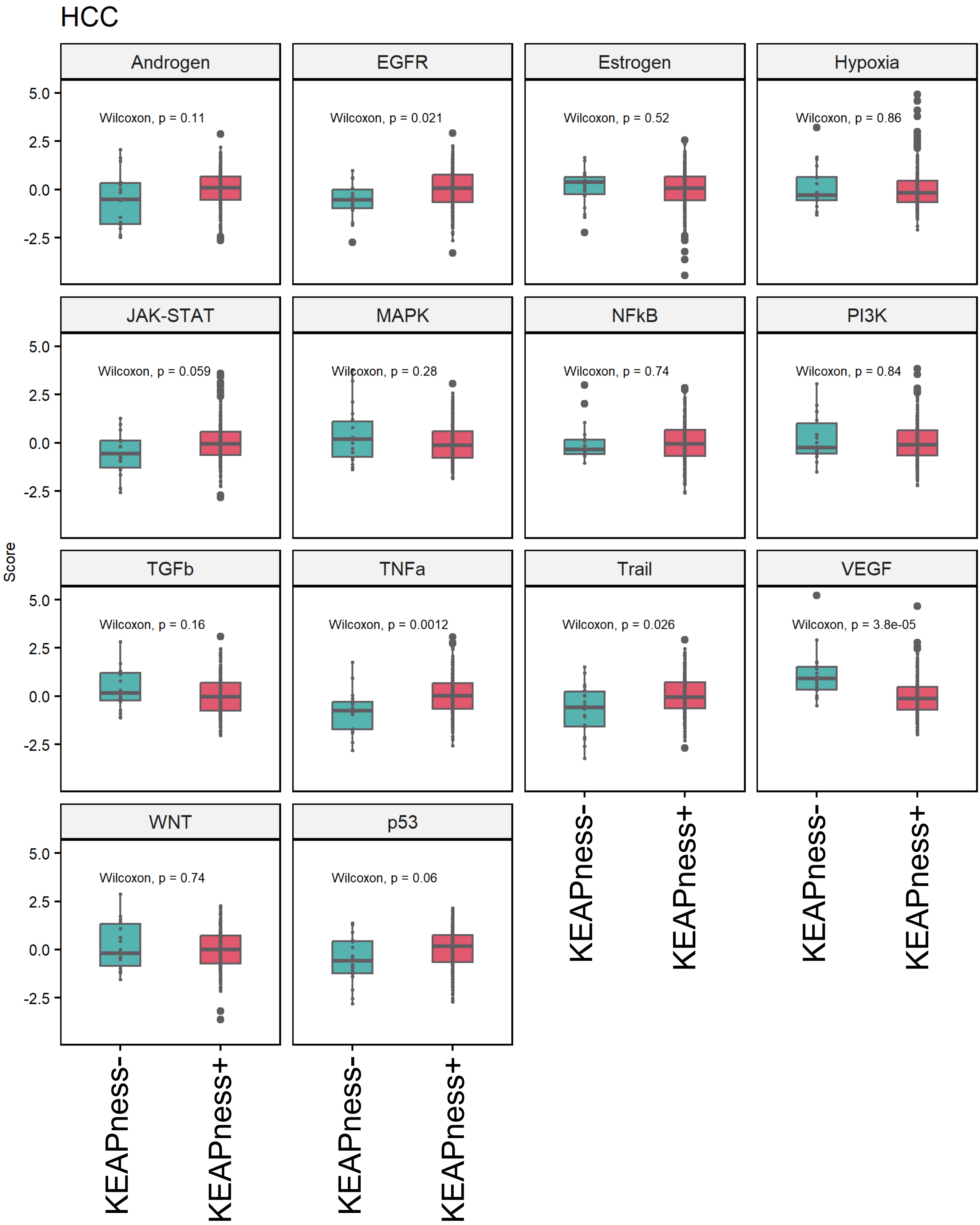

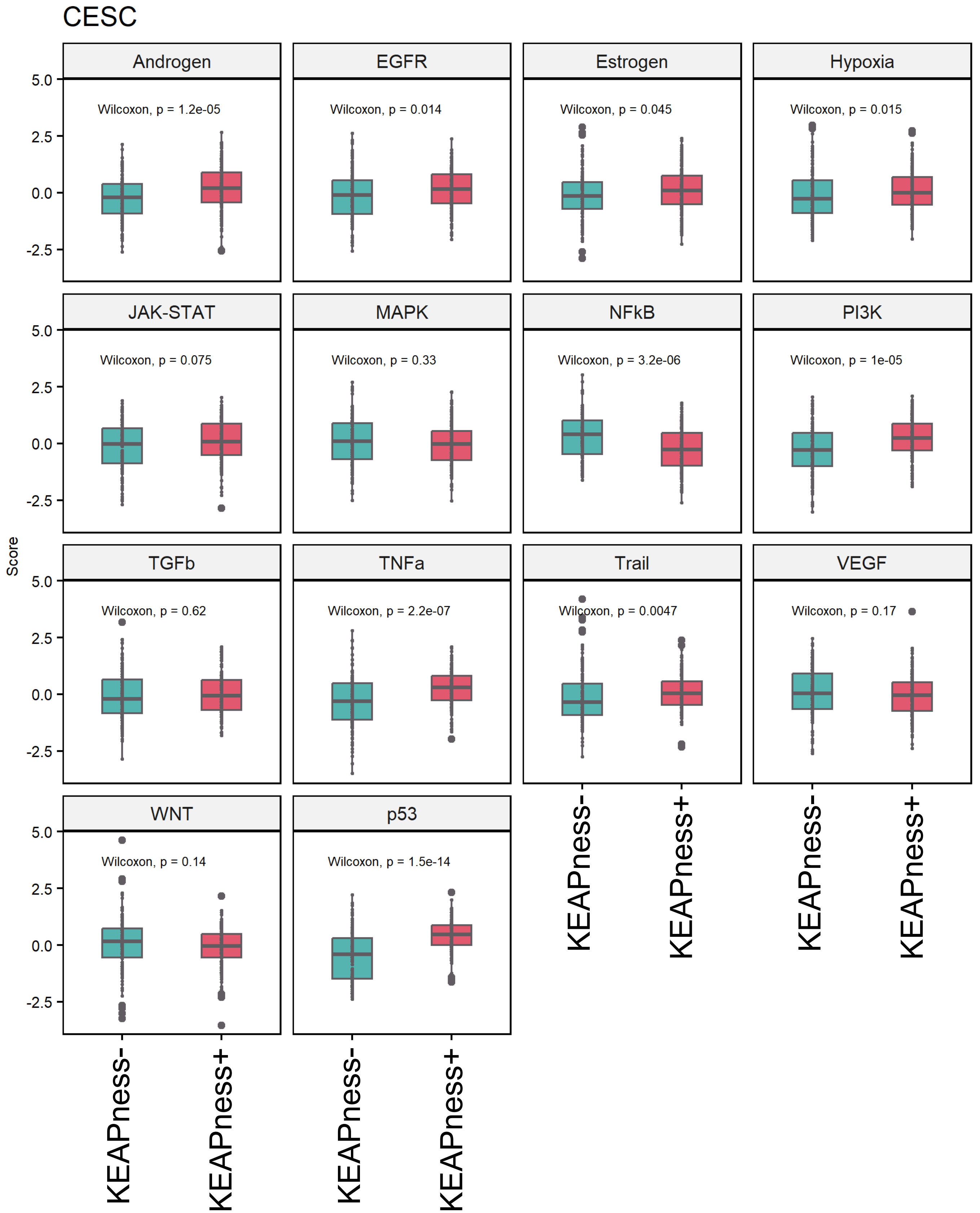

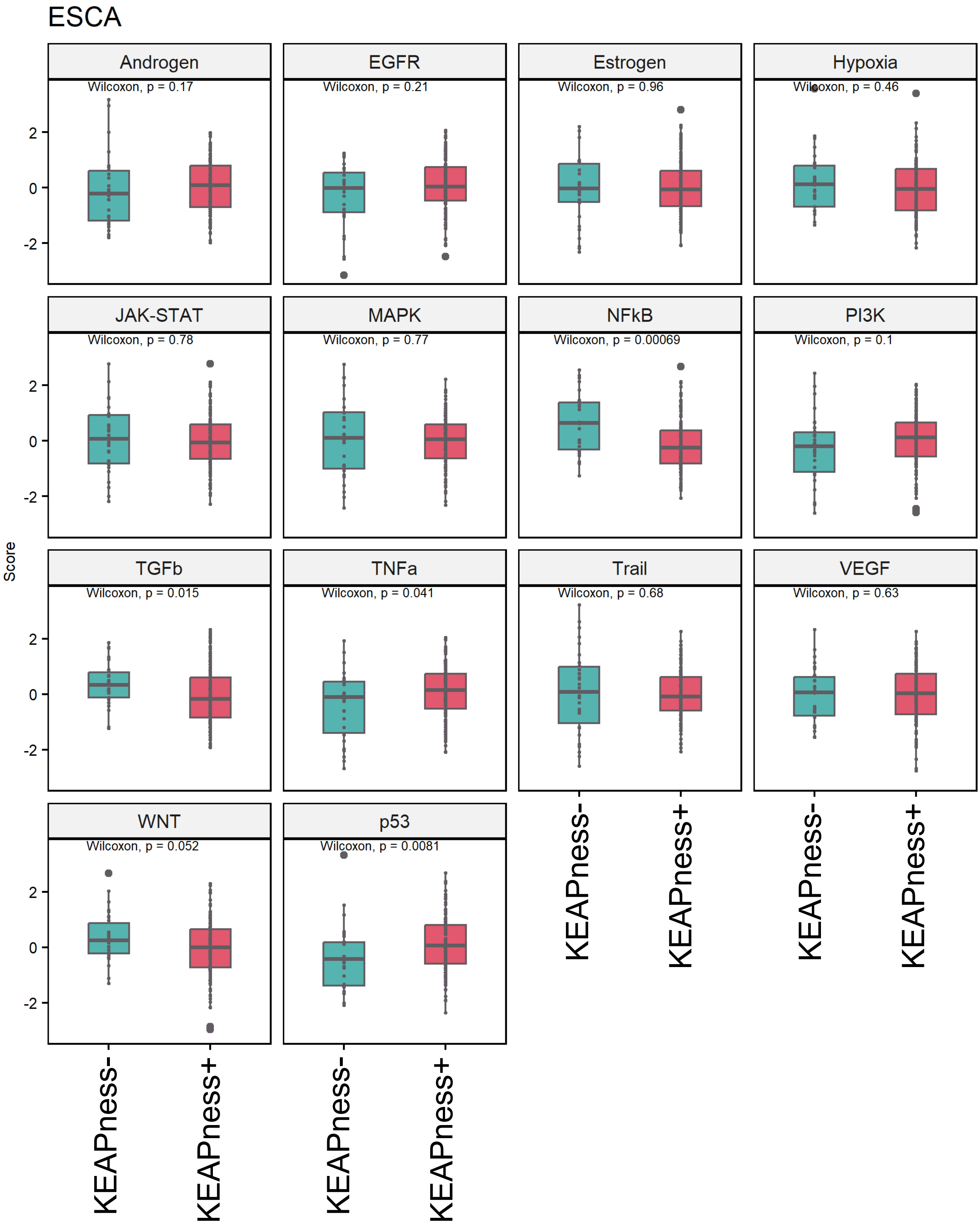

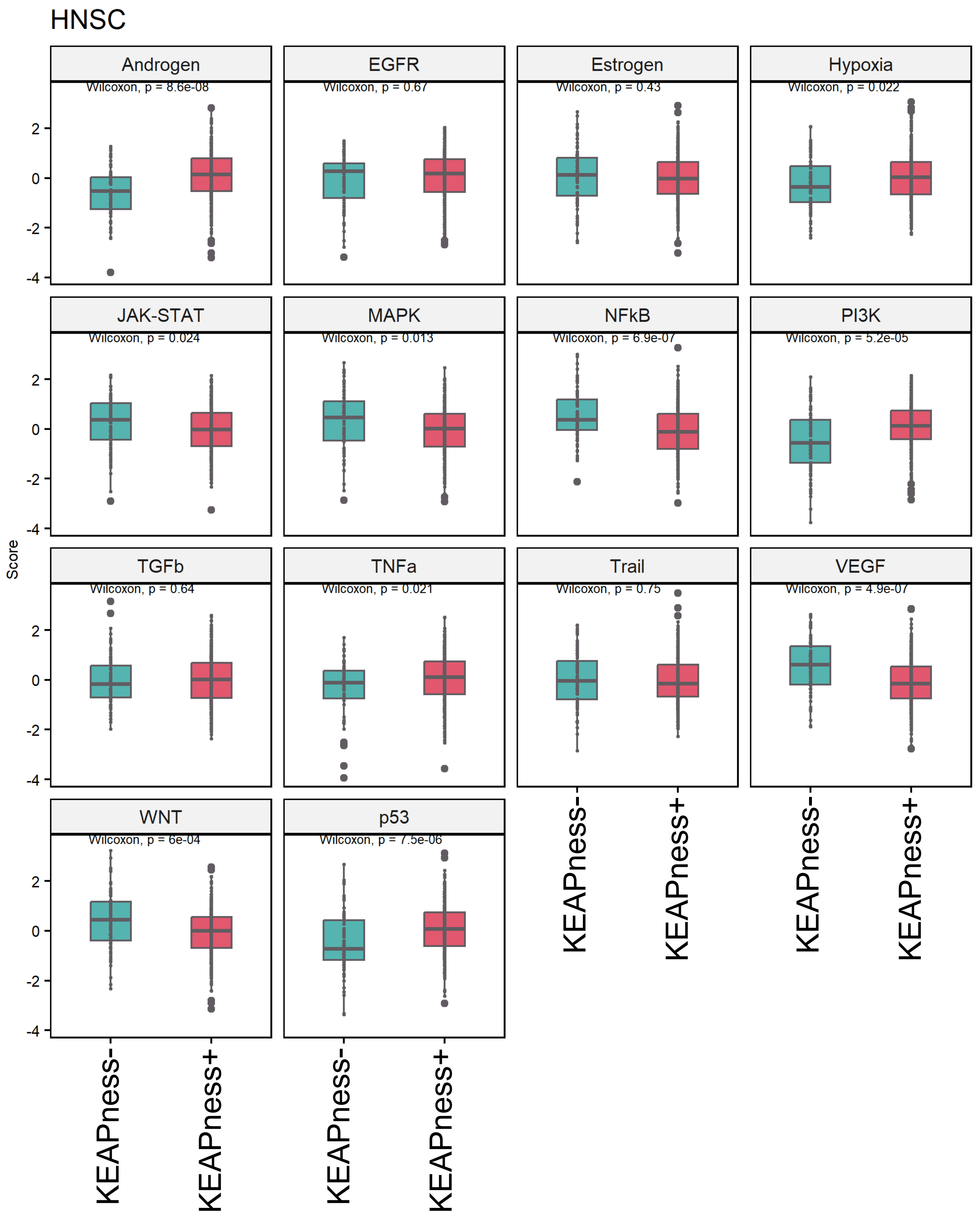

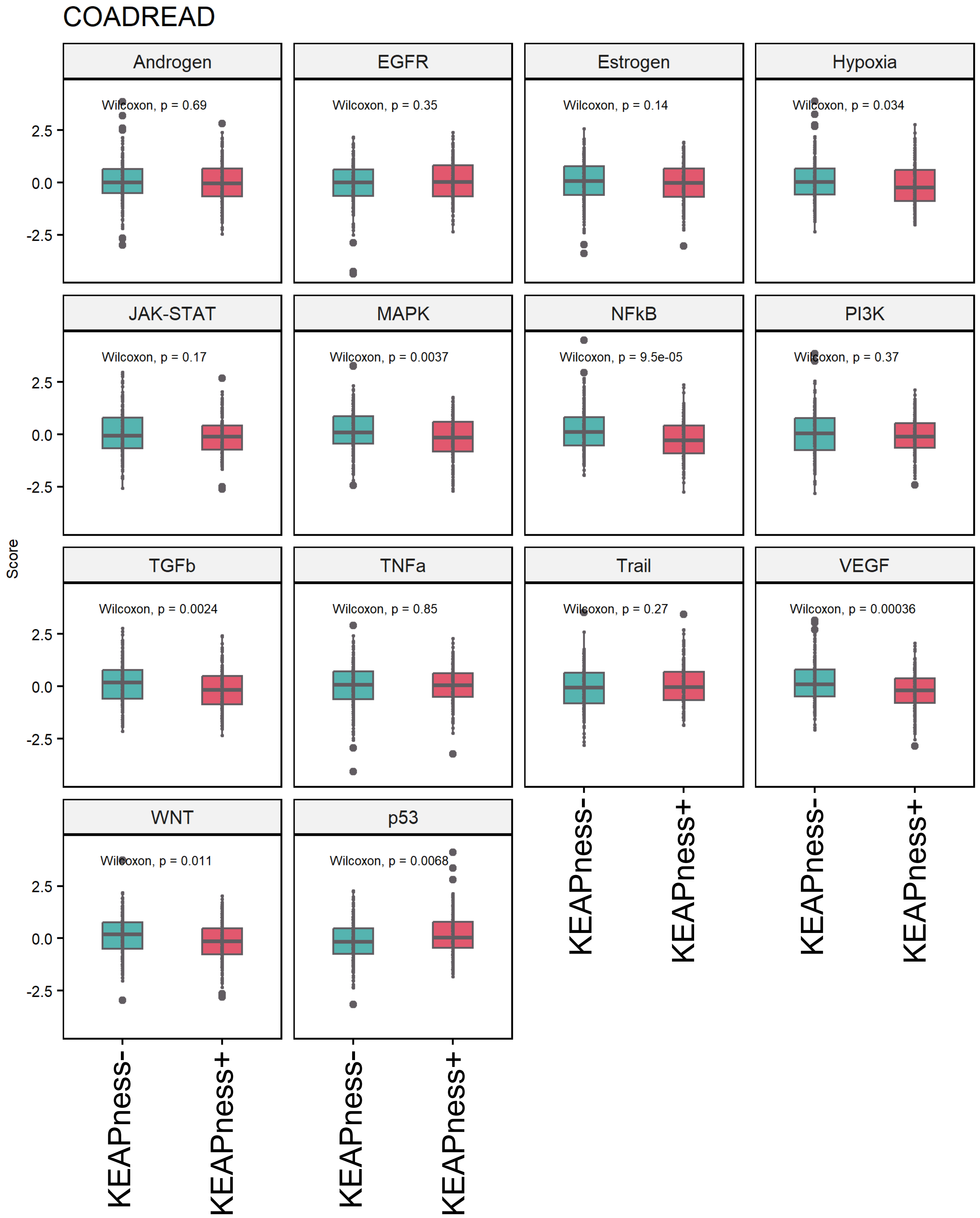

### Supplementary Figure 3

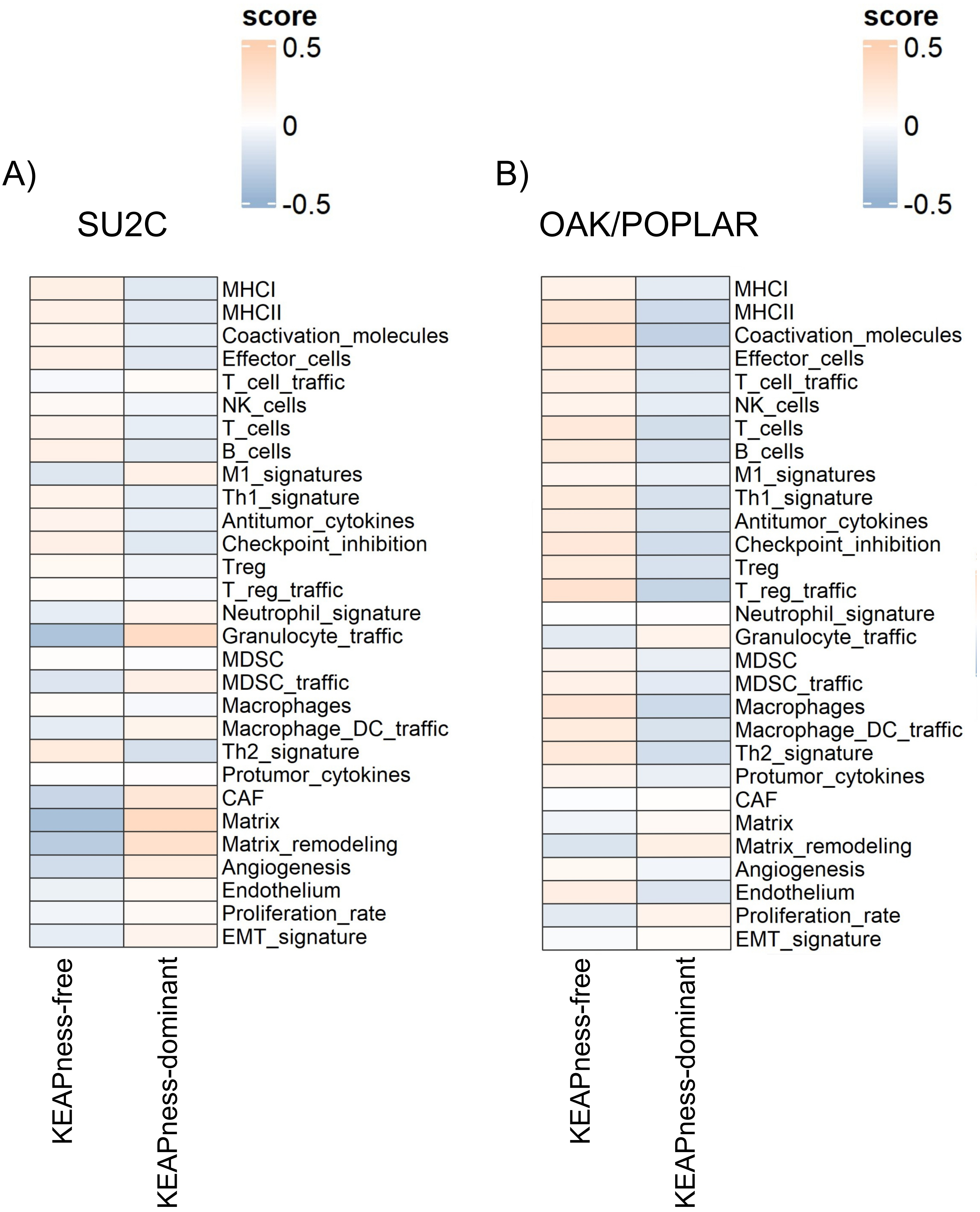

### Supplementary Figure 4

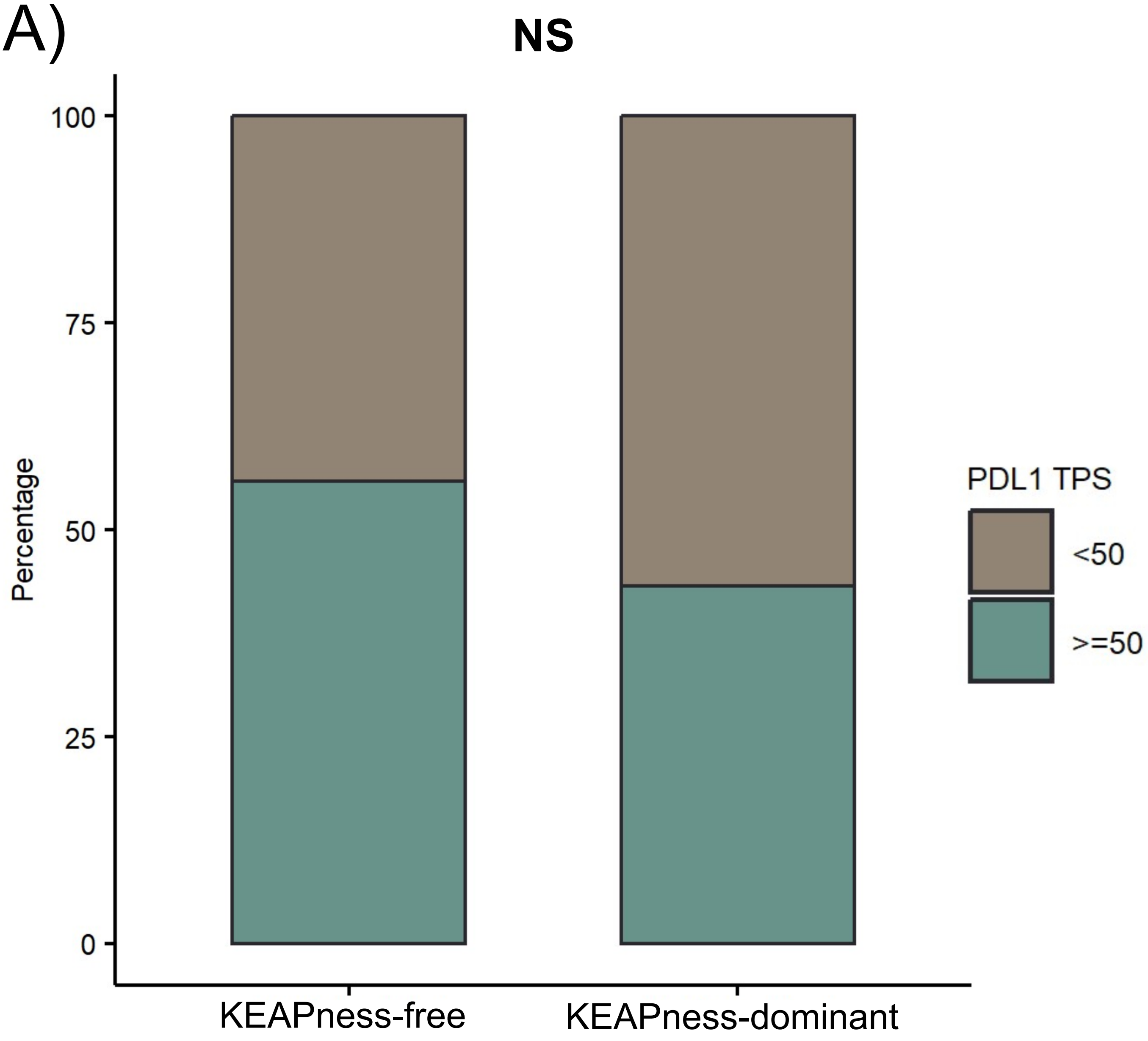

### Supplementary Figure 5

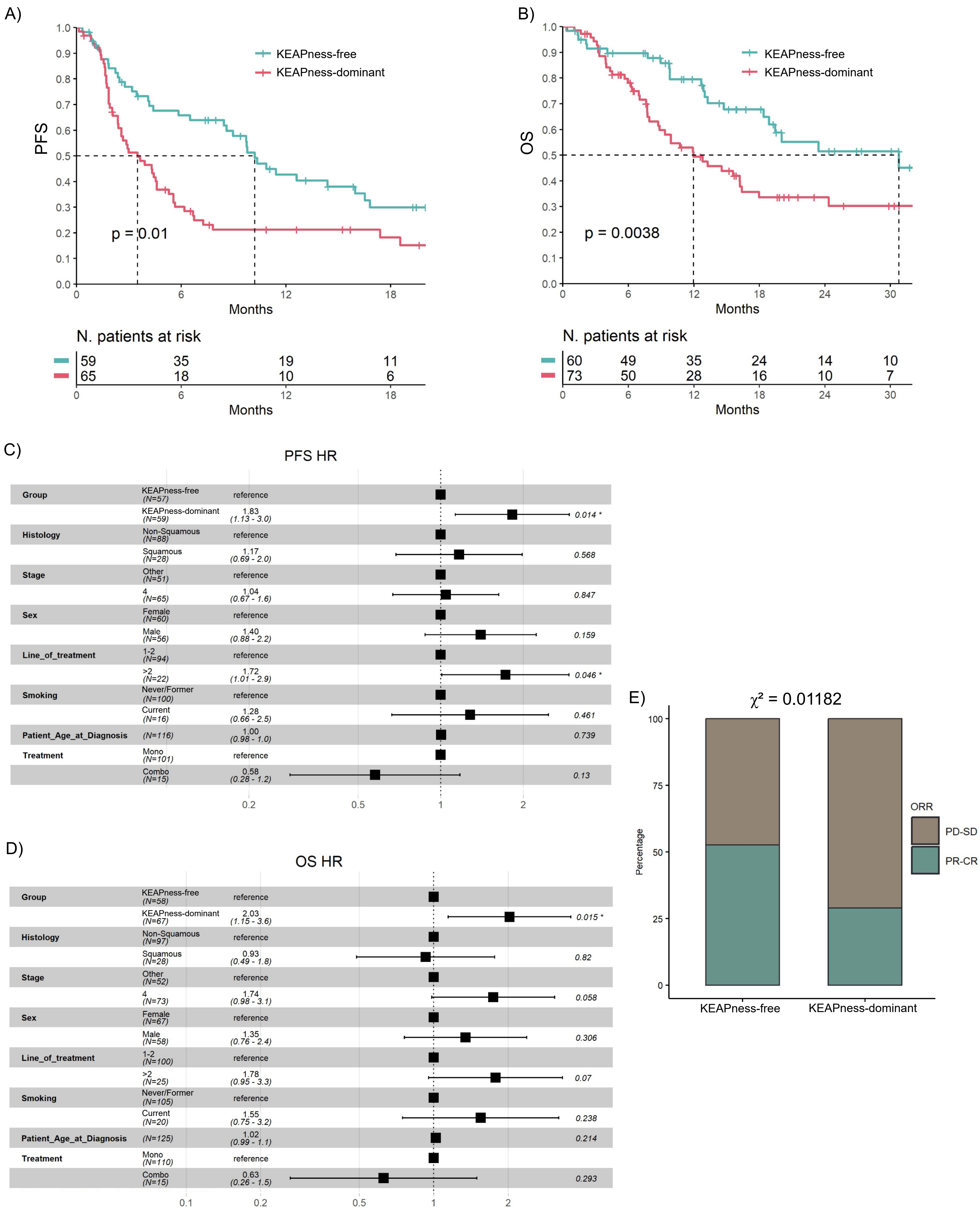

### Supplementary Figure 6

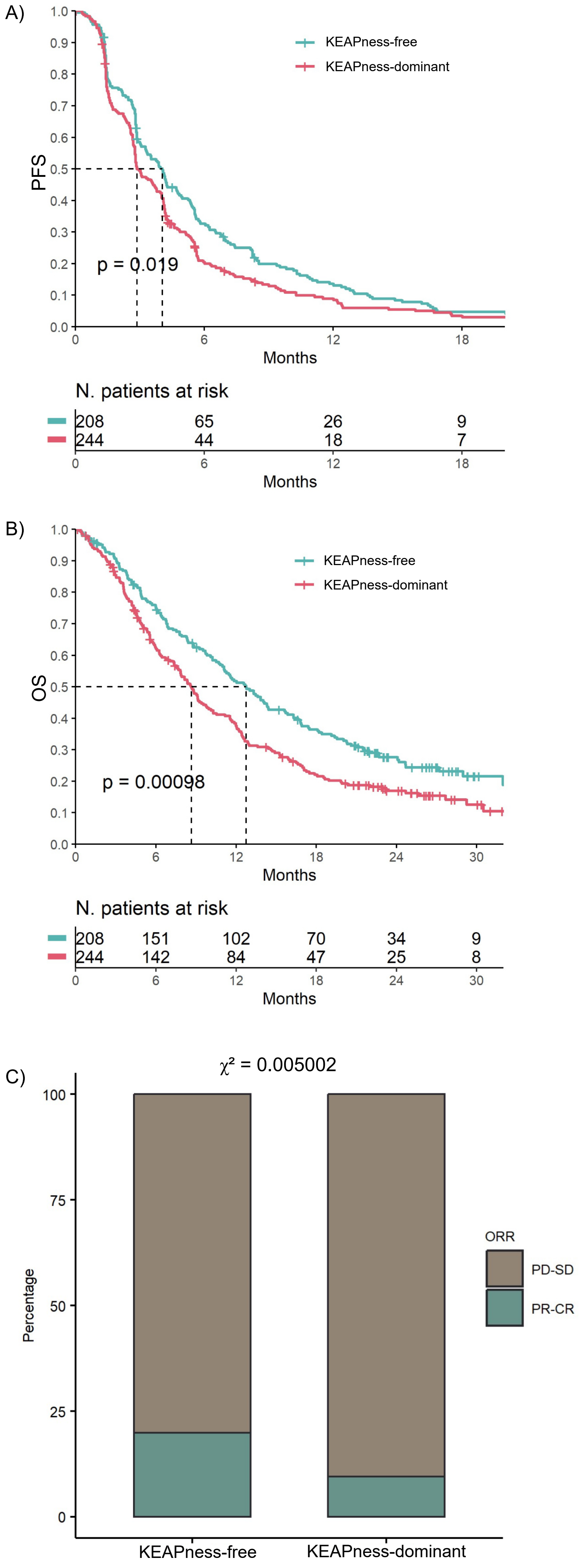

### Supplementary Figure 7

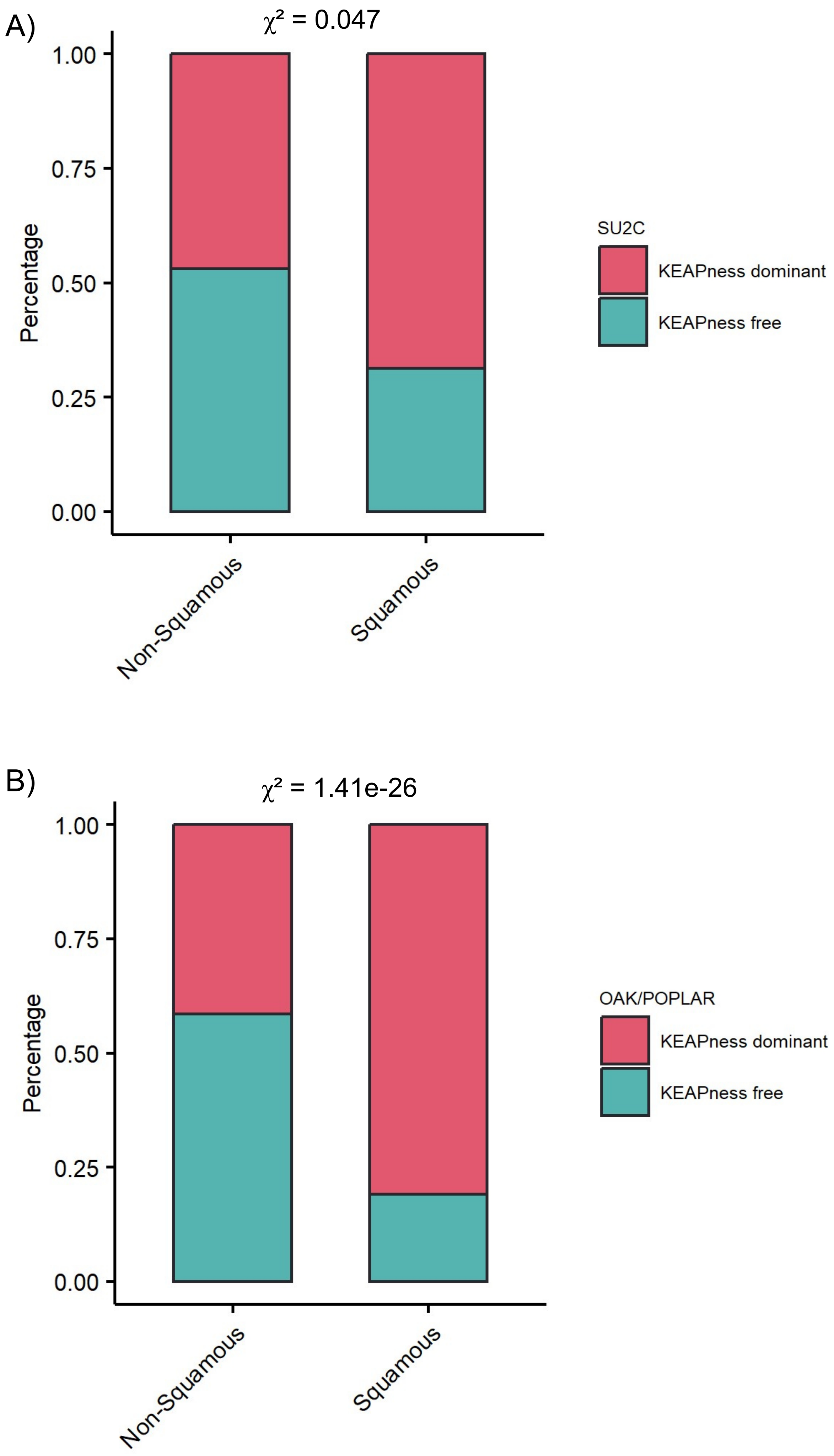

### Supplementary Figure 8

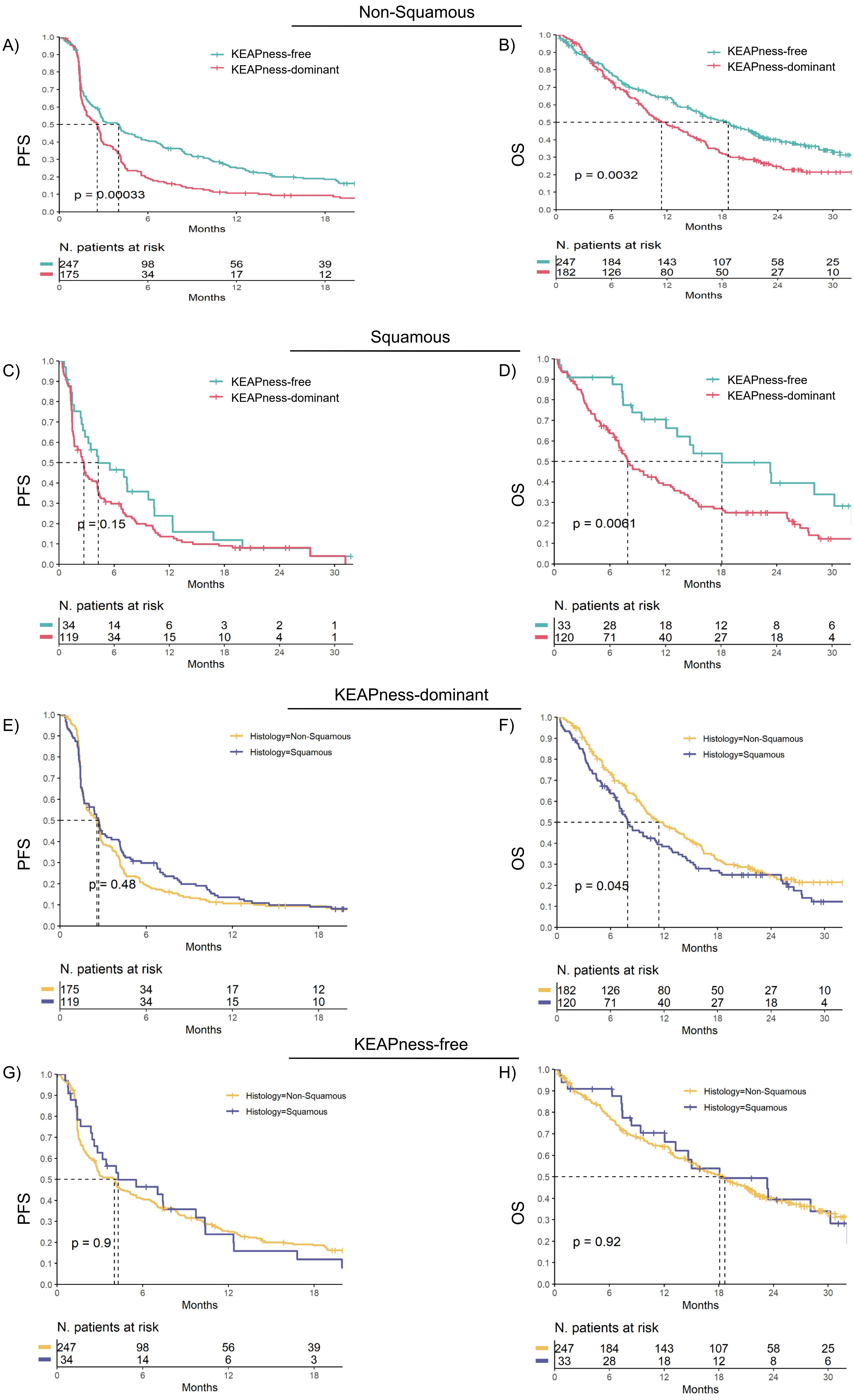
